# Inequalities in antenatal care coverage and quality: an analysis from 63 low and middle-income countries using the ANCq content-qualified coverage indicator

**DOI:** 10.1101/2020.11.11.20230102

**Authors:** Luisa Arroyave, Ghada E Saad, Cesar G Victora, Aluisio J D Barros

## Abstract

**Objective:** to conduct a global analysis of socioeconomic inequalities in antenatal care (ANC) using national surveys from low- and middle-income countries.

**Methods:** ANC was measured using the ANCq, a novel content-qualified ANC coverage indicator, created and validated using national surveys, based upon contact with the health services and content of care received. We performed stratified analysis to explore the socioeconomic inequalities in ANCq. We also estimated the slope index of inequality, which measures the difference in coverage along the wealth spectrum.

**Results:** We analyzed 63 national surveys carried out from 2010 to 2017. There were large inequalities between and within countries. Higher ANCq scores were observed among women living in urban areas, with secondary or more level of education, belonging to wealthier families and with higher empowerment in nearly all countries. Countries with higher ANCq mean presented lower inequalities; while countries with average ANCq scores presented wide range of inequality, with some managing to achieve very low inequality.

**Conclusions:** Despite all efforts in ANC programs, important inequalities in coverage and quality of ANC services persist. If maternal and child mortality Sustainable Development Goals are to be achieved, those gaps we documented must be bridged.

## Introduction

Improving maternal and reproductive health remains a challenge in low and middle-income countries (LMICs), where the most vulnerable women have limited or no access to health services, and poor quality care, therefore presenting the worst maternal health outcomes.^1,2^ Good quality antenatal care (ANC) helps to reduce adverse maternal and newborn outcomes.^3–6^ However, measuring it has been a challenge, mainly due to lack of information from household surveys about content of care and that can be applied for a large number of countries. Traditionally, surveys record the number of antenatal care visits, the provider of care and a few interventions, such as measuring blood pressure or collecting samples of urine and blood. It is a small part of what ANC is expected to offer, and the information collected varies widely from surveys in different countries.^7^

The ANCq – content-qualified ANC coverage indicator^8^ – was proposed as a new indicator that combines a set of key aspects of contact with services and content of care. In contrast to most of the existing ANC indicators, ANCq is calculated as a score giving an idea of level of adequacy, and also considering all pregnant woman in need of ANC and not only those who had at least one visit. It was created and validated based on national surveys from 63 LMICs, showing wide variation in the ANCq mean scores between countries and world regions. Thus, it is important to explore the inequalities related to ANC, also considering the evidence suggested substantial inequalities in maternal and child health, and the effects it may have on the lives of people.^1,2,9^ Inequalities in health care access and services are considered a multidimensional issue that weakens and delays overall country development and progress, particularly in LMICs due to low socioeconomic levels and lack of opportunities for women’s empowerment.^9^

This paper presents a global analysis of socioeconomic inequalities in ANC, using the ANCq indicator. Using data from nationally representative household surveys carried out in LMICs, inequalities in terms of wealth, place of residence, woman’s age and education, sex of the child, and woman’s empowerment were explored.

## Methods

This study was based on nationally representative health surveys, including Demographic and Health Survey (DHS) and Multiple Indicator Cluster Survey (MICS). Both types of surveys use standardized data collection procedures, making the results comparable across surveys and countries.^10–12^

The analysis included the latest DHS or MICS survey from 63 LMICs carried out from 2010 to 2017, with information that was enough for the calculation of the ANCq – content qualified ANC indicator – used in our analysis to measure ANC.

ANCq is a novel survey-based ANC indicator calculated as a score, composed of seven variables which add points to the score: first visit in the first trimester of pregnancy (1 point), at least one visit with a skilled provider (2 points), total number of visits (1 point for 1-3 visits, 2 points for 4-7 visits, and 3 points for 8 or more visits), blood pressure measured (1 point), blood sample collected (1 point), urine sample collected (1 point), and receiving at least two shots of tetanus toxoid (1 point). Thus, the ANCq score varies from zero, for women with no ANC, to 10 points, for women getting top points for each item. ANCq was validated using a convergent validation exercise exploring the association with neonatal mortality, where higher scores of ANCq were associated with lower neonatal mortality. Full details on the construction of the indicator and its validity are presented elsewhere.^8^

In this paper, we performed stratified analysis to explore inequalities in the ANCq indicator by wealth, area of residence, woman’s age and education, woman’s empowerment, and sex of the child. Each stratification variable is defined below:^13^

- Place of residence: urban or rural based on criteria defined by each country.
- Woman’s age: three groups of age, at the time the child was born: 15-19, 20-34, 35-49 years.
- Woman’s education: three categories: none (no formal education); primary (any primary education, including completed primary education) and secondary or higher (any secondary education, including complete secondary; this category also includes women with partial or full higher education).
- Sex of the child: female or male.
- Wealth quintiles: based on an asset index obtained from information on characteristics of the building materials, household assets, presence of electricity, water supply and sanitary facilities, amongst other.^14,15^ Because relevant assets may vary in urban and rural households, separate principal component analyses are carried out in each area. The resulting scores are combined into a single one using a scaling procedure to allow comparability between urban and rural households. The sample is divided into quintiles ranging from quintile 1 representing approximately the poorest 20% of women in the surveys sample and quintile 5, that represents the wealthiest 20%.^16^
- Woman’s empowerment: measured using the three domains of the Survey-based Women’s emPowERment (SWPER) index: attitude to violence, social independence and decision making. The SWPER is based on 14 questions related to the women’s opinion on whether beating the wife is justified in some situations, involvement in household decisions, women’s education, access to information, age at marriage and first child, and difference in age and education between the woman and her husband.^17,18^

For woman’s empowerment and sex of the child, we only used DHS because we can directly link the relevant datasets needed. Woman’s empowerment was calculated only for those surveys with available information to create the SWPER.

ANCq estimates for countries are presented with their 95% confidence interval (95% CI), for each defined stratification variable. Equiplots are presented to visually show the inequalities, between and within countries. Countries were grouped according to UNICEF regions. Regional estimates were weighted by the size of population of women (15-49 years) obtained from World Bank Population Estimates and Projections^19^ in the year when each survey was carried out.

From our initial set of countries, we selected 12 to further explore the coverage level of each component of the ANCq so that we could better understand which are the bottlenecks and which are the component indicators that achieve high coverage for most groups. Three countries were selected from each of the quadrants in Figure 2, representing countries with high inequality and low ANCq score, high inequality and high score, low inequality and low score, and low inequality and high score.

**Figure 1.**
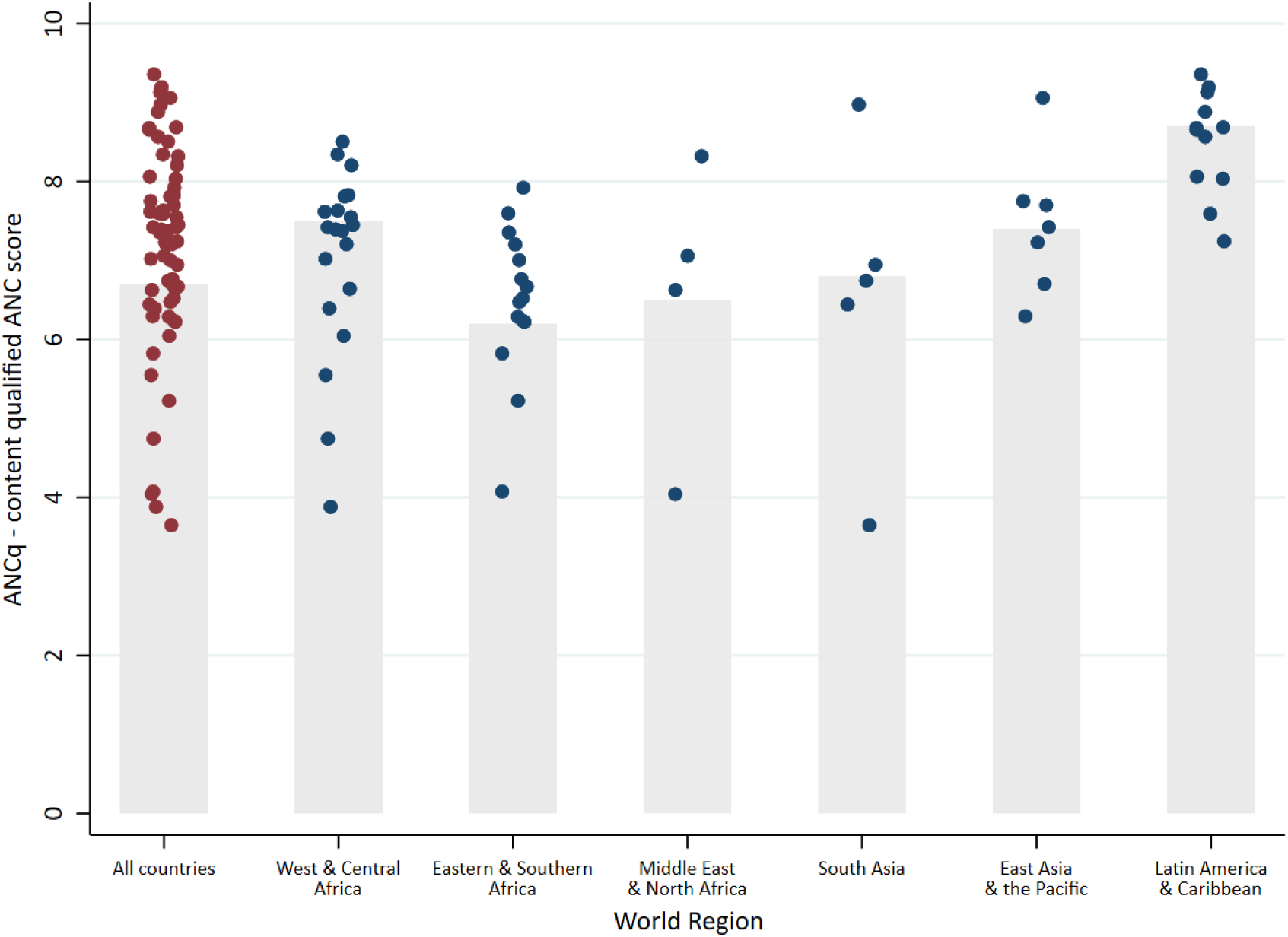
ANCq means for 63 LMICs, by UNICEF regions. The gray bars show the region weighted median for the countries with data. Source: DHS and MICS, 2010-2017.

**Figure 2.**
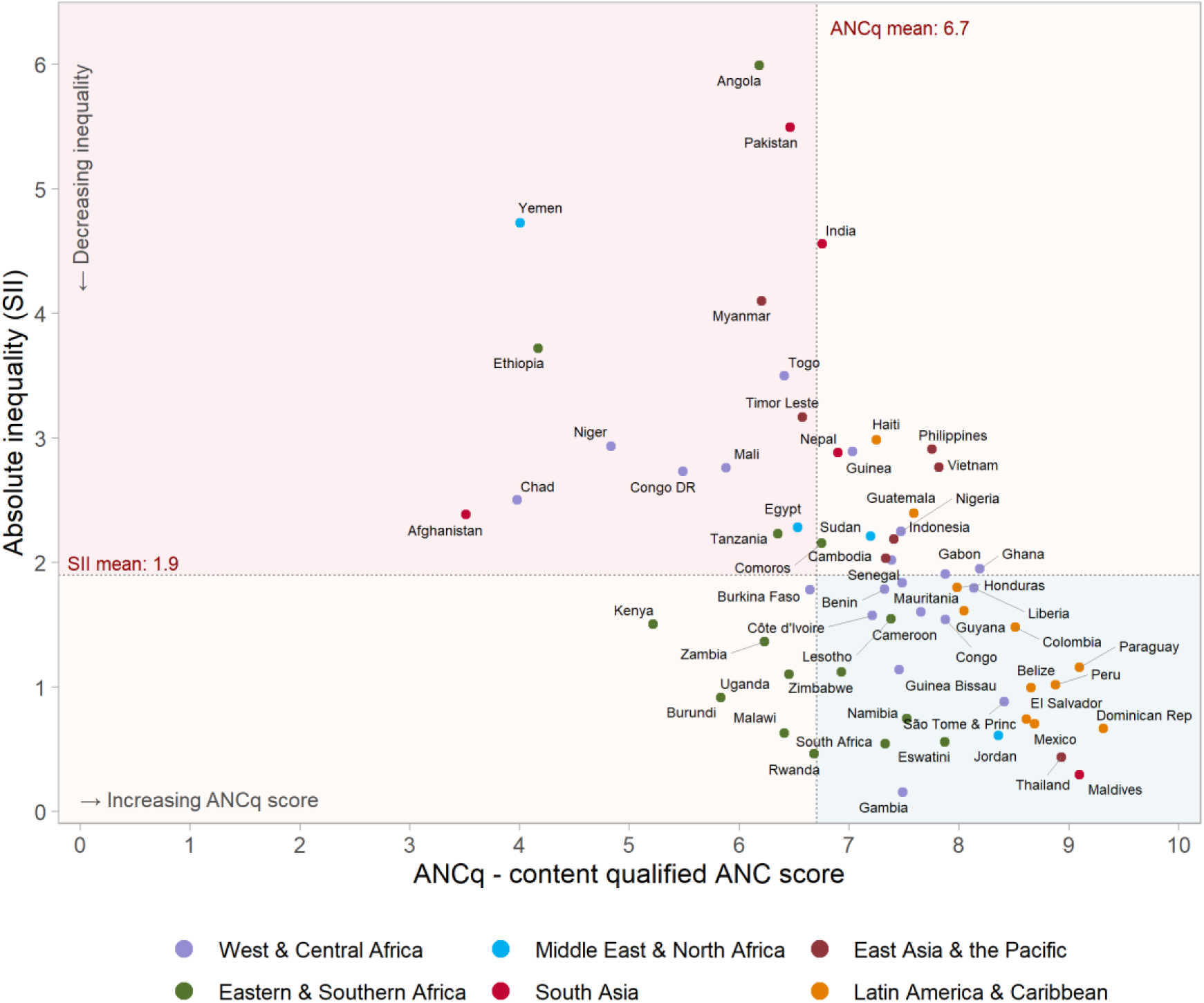
Slope Index of Inequality (absolute inequalities) of ANCq score for 63 LMICs. Source: DHS and MICS, 2010-2017.

Absolute inequality was measured with the slope index of inequality (SII), derived through a linear regression model where the outcome was the ANCq. SII “represents the absolute difference in the fitted value of the indicator between the highest and the lowest values of the socioeconomic indicator rank”.^20^ The SII was also estimated for each ANCq component in the 12 selected countries to explore low coverage and high inequalities for the component indicator. In this case we used a logistic regression model given the components are binary variables, except for number of visits.^20^

The analyses were performed using Stata (StataCorp. 2019. Stata Statistical Software: Release 16. College Station, TX: StataCorp LLC), always considering the survey design (clustering and sampling weights).

The study was based on an anonymized publicly available data. Ethical clearance was done by each of the institutions responsible for carrying out the original surveys.

## Results

We analyzed 63 national surveys with available data to calculate the ANCq indicator. Table 1 shows the list of surveys grouped by the UNICEF regions (no country from Europe and Central Asia had enough data), with the ANCq mean and respective SII.

**Table 1.**
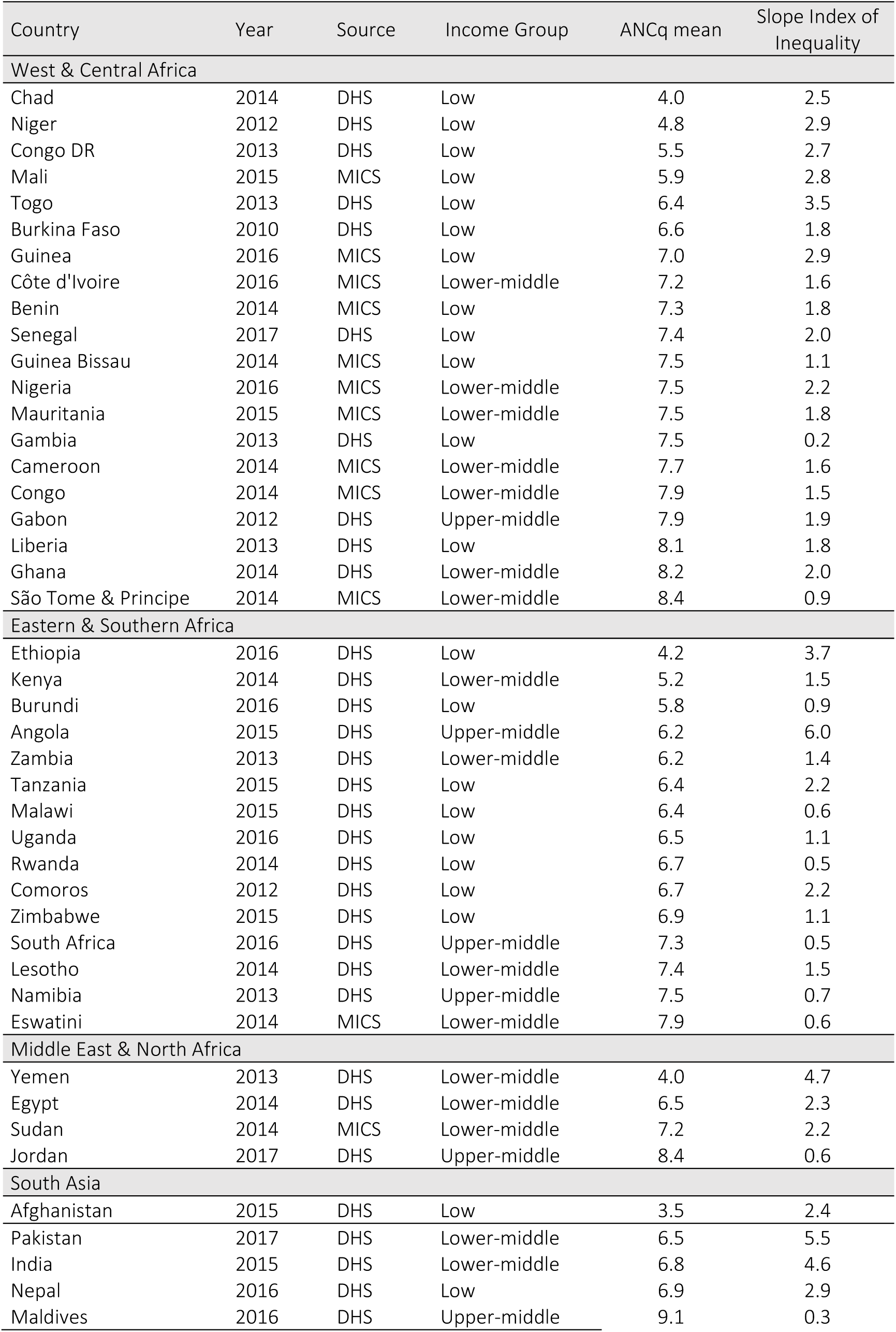
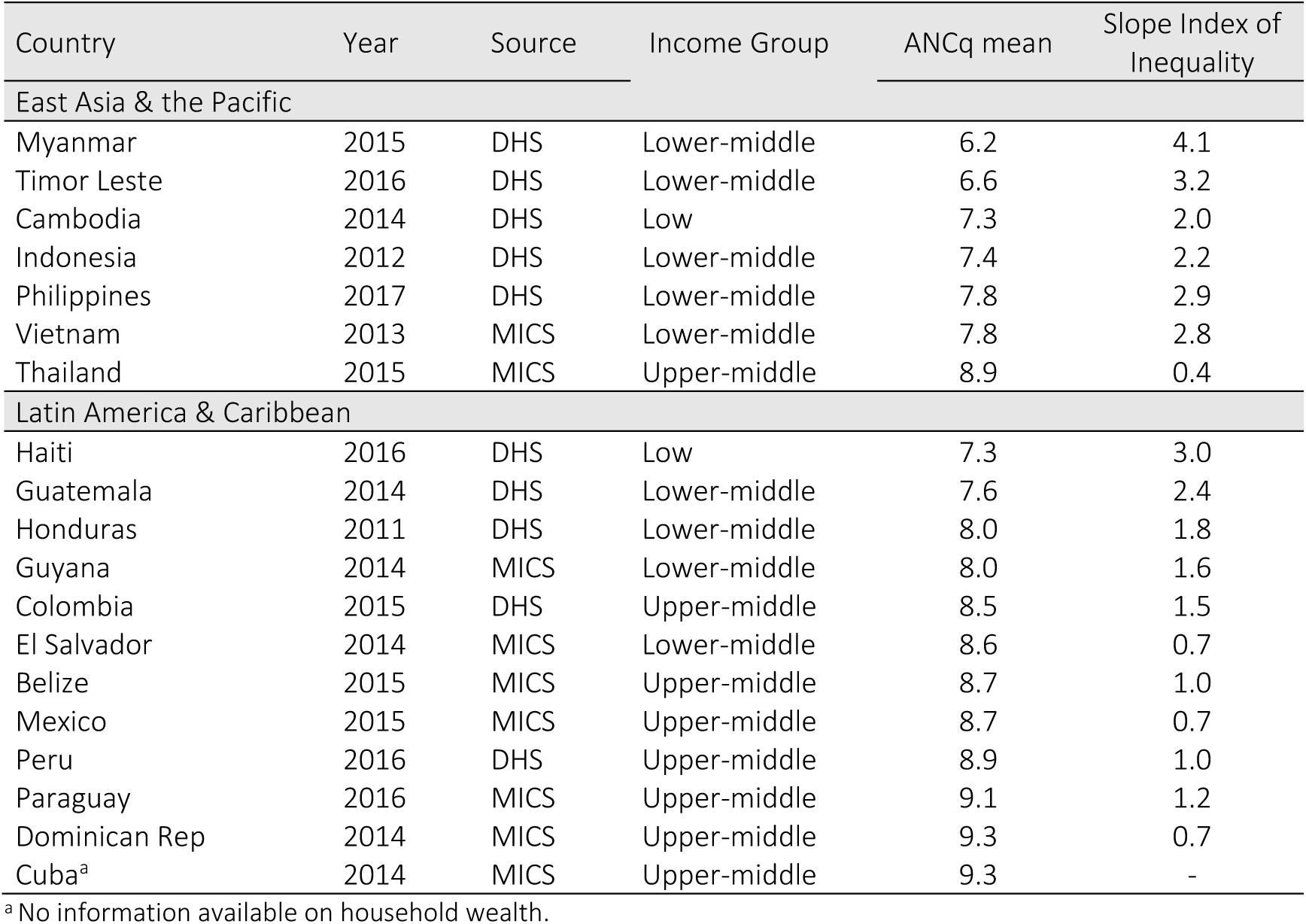
ANCq mean and Slope Index of Inequality (absolute inequalities) of ANCq score for 63 LMICs, sorted by ANCq mean within UNICEF region. Source: DHS and MICS, 2010-2017.

| Country | Year | Source | Income Group | ANCq mean | Slope Index of Inequality |
| --- | --- | --- | --- | --- | --- |
| West & Central Africa |  |  |  |  |  |
| Chad | 2014 | DHS | Low | 4.0 | 2.5 |
| Niger | 2012 | DHS | Low | 4.8 | 2.9 |
| Congo DR | 2013 | DHS | Low | 5.5 | 2.7 |
| Mali | 2015 | MICS | Low | 5.9 | 2.8 |
| Togo | 2013 | DHS | Low | 6.4 | 3.5 |
| Burkina Faso | 2010 | DHS | Low | 6.6 | 1.8 |
| Guinea | 2016 | MICS | Low | 7.0 | 2.9 |
| Côte d'Ivoire | 2016 | MICS | Lower-middle | 7.2 | 1.6 |
| Benin | 2014 | MICS | Low | 7.3 | 1.8 |
| Senegal | 2017 | DHS | Low | 7.4 | 2.0 |
| Guinea Bissau | 2014 | MICS | Low | 7.5 | 1.1 |
| Nigeria | 2016 | MICS | Lower-middle | 7.5 | 2.2 |
| Mauritania | 2015 | MICS | Lower-middle | 7.5 | 1.8 |
| Gambia | 2013 | DHS | Low | 7.5 | 0.2 |
| Cameroon | 2014 | MICS | Lower-middle | 7.7 | 1.6 |
| Congo | 2014 | MICS | Lower-middle | 7.9 | 1.5 |
| Gabon | 2012 | DHS | Upper-middle | 7.9 | 1.9 |
| Liberia | 2013 | DHS | Low | 8.1 | 1.8 |
| Ghana | 2014 | DHS | Lower-middle | 8.2 | 2.0 |
| São Tome & Príncipe | 2014 | MICS | Lower-middle | 8.4 | 0.9 |
| Eastern & Southern Africa |  |  |  |  |  |
| Ethiopia | 2016 | DHS | Low | 4.2 | 3.7 |
| Kenya | 2014 | DHS | Lower-middle | 5.2 | 1.5 |
| Burundi | 2016 | DHS | Low | 5.8 | 0.9 |
| Angola | 2015 | DHS | Upper-middle | 6.2 | 6.0 |
| Zambia | 2013 | DHS | Lower-middle | 6.2 | 1.4 |
| Tanzania | 2015 | DHS | Low | 6.4 | 2.2 |
| Malawi | 2015 | DHS | Low | 6.4 | 0.6 |
| Uganda | 2016 | DHS | Low | 6.5 | 1.1 |
| Rwanda | 2014 | DHS | Low | 6.7 | 0.5 |
| Comoros | 2012 | DHS | Low | 6.7 | 2.2 |
| Zimbabwe | 2015 | DHS | Low | 6.9 | 1.1 |
| South Africa | 2016 | DHS | Upper-middle | 7.3 | 0.5 |
| Lesotho | 2014 | DHS | Lower-middle | 7.4 | 1.5 |
| Namibia | 2013 | DHS | Upper-middle | 7.5 | 0.7 |
| Eswatini | 2014 | MICS | Lower-middle | 7.9 | 0.6 |
| Middle East & North Africa |  |  |  |  |  |
| Yemen | 2013 | DHS | Lower-middle | 4.0 | 4.7 |
| Egypt | 2014 | DHS | Lower-middle | 6.5 | 2.3 |
| Sudan | 2014 | MICS | Lower-middle | 7.2 | 2.2 |
| Jordan | 2017 | DHS | Upper-middle | 8.4 | 0.6 |
| South Asia |  |  |  |  |  |
| Afghanistan | 2015 | DHS | Low | 3.5 | 2.4 |
| Pakistan | 2017 | DHS | Lower-middle | 6.5 | 5.5 |
| India | 2015 | DHS | Lower-middle | 6.8 | 4.6 |
| Nepal | 2016 | DHS | Low | 6.9 | 2.9 |
| Maldives | 2016 | DHS | Upper-middle | 9.1 | 0.3 |
| East Asia & the Pacific |  |  |  |  |  |
| Myanmar | 2015 | DHS | Lower-middle | 6.2 | 4.1 |
| Timor Leste | 2016 | DHS | Lower-middle | 6.6 | 3.2 |
| Cambodia | 2014 | DHS | Low | 7.3 | 2.0 |
| Indonesia | 2012 | DHS | Lower-middle | 7.4 | 2.2 |
| Philippines | 2017 | DHS | Lower-middle | 7.8 | 2.9 |
| Vietnam | 2013 | MICS | Lower-middle | 7.8 | 2.8 |
| Thailand | 2015 | MICS | Upper-middle | 8.9 | 0.4 |
| Latin America & Caribbean |  |  |  |  |  |
| Haiti | 2016 | DHS | Low | 7.3 | 3.0 |
| Guatemala | 2014 | DHS | Lower-middle | 7.6 | 2.4 |
| Honduras | 2011 | DHS | Lower-middle | 8.0 | 1.8 |
| Guyana | 2014 | MICS | Lower-middle | 8.0 | 1.6 |
| Colombia | 2015 | DHS | Upper-middle | 8.5 | 1.5 |
| El Salvador | 2014 | MICS | Lower-middle | 8.6 | 0.7 |
| Belize | 2015 | MICS | Upper-middle | 8.7 | 1.0 |
| Mexico | 2015 | MICS | Upper-middle | 8.7 | 0.7 |
| Peru | 2016 | DHS | Upper-middle | 8.9 | 1.0 |
| Paraguay | 2016 | MICS | Upper-middle | 9.1 | 1.2 |
| Dominican Rep | 2014 | MICS | Upper-middle | 9.3 | 0.7 |
| Cuba<sup>a</sup> | 2014 | MICS | Upper-middle | 9.3 | - |

<sup>a</sup> No information available on household wealth.

Wide variation in ANCq was observed between and within the regions. Figure 1 shows the average scores of ANCq for each country in each region (blue dots), and the ANCq median for regions (gray bar). South Asia, and Middle East and North Africa were the regions with the widest spread of ANCq. In South Asia, the ANCq ranged from 3.5 in Afghanistan to 9.1 in the Maldives (Table 1). The Latin America and Caribbean region presented the lowest between country inequality and the highest ANCq median score (8.6) (Figure 1). ANCq in Latin American countries ranged between 7.3 in Haiti to 9.3 in Cuba and the Dominican Republic (Table 1).

Large within country wealth-related inequalities were observed in several countries. Angola, Pakistan, Yemen, India, Myanmar, Ethiopia, and Togo were, in decreasing order, the countries with the highest SII, ranging from 6.0 to 3.5 (Table 1). That is, the difference between the top and bottom of the wealth scale in these countries were as large as six ANCq points.

In general, countries with a higher mean ANCq presented lower SII values. The Pearson correlation between the two indicators was −0.52 (p<0.001). Figure 2 shows the average scores for ANCq plotted against the SII. Countries located in the upper-left quadrant are the ones that stand out with low ANCq scores and high inequality, while those in the lower-right quadrant are the best positioned presenting higher ANCq and lower SII. Maldives and Thailand are the best positioned countries in this group. It is also easy from Figure 2 to depict countries with the lowest or highest levels of inequality.

The mean ANCq for each wealth quintile is shown in Figure 3, where quintiles are nearly always ordered from Q1 to Q5 indicating a very systematic monotonic increase of ANCq with wealth. Several of the countries with the highest inequalities had their richest quintile positioned close to the countries with the best ANCq scores. This is the case of Angola, Pakistan, India, and Myanmar. On the other extreme, the poorest groups in these countries are among those with the lowest scores.

**Figure 3.**
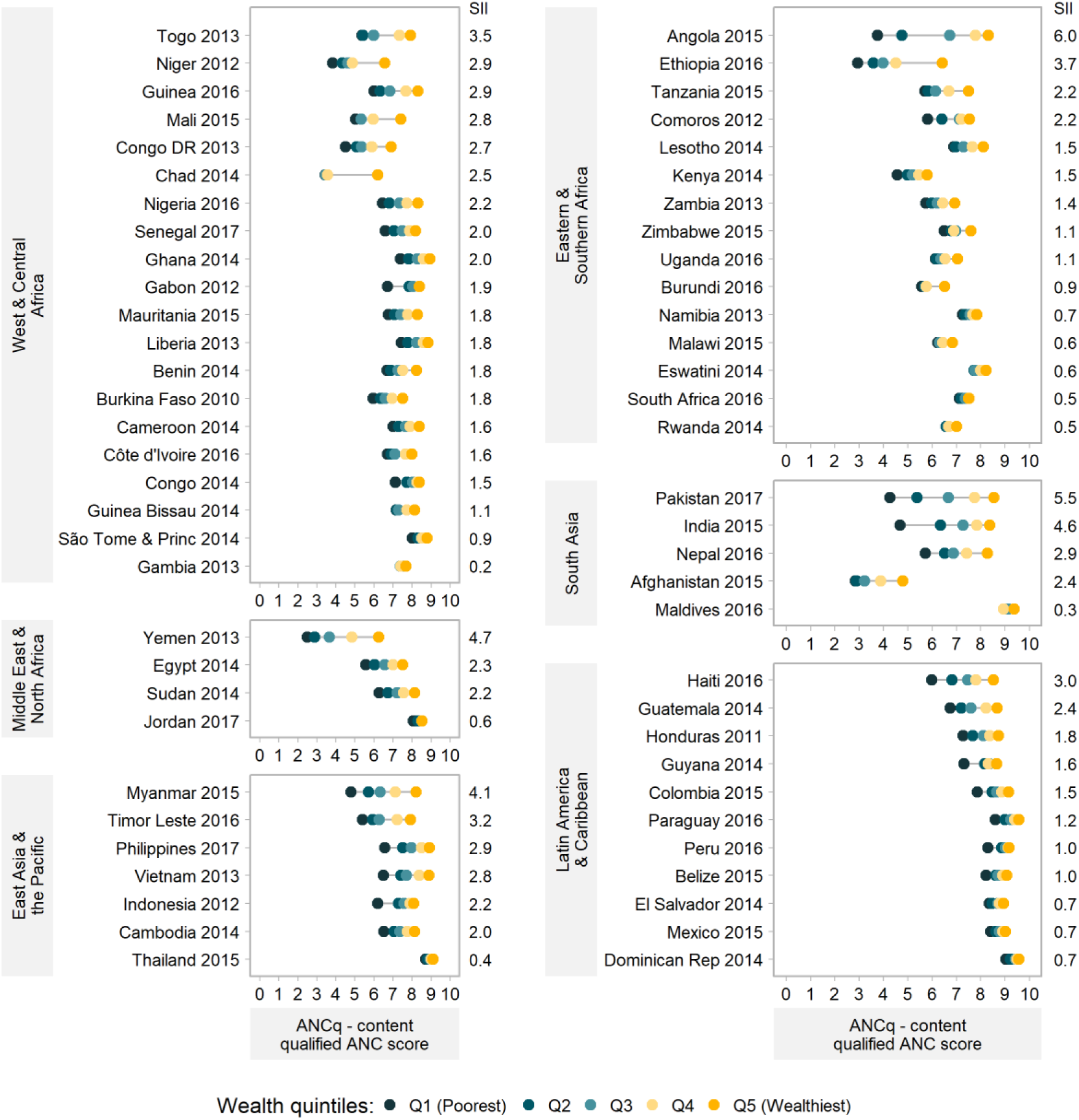
Equiplots of ANCq score by wealth quintiles. Countries are ordered by decreasing inequality (SII) in each region. Source: DHS and MICS, 2010-2017.

The coverage for each component indicator of the ANCq for the richest and poorest quintiles along with their respective SII for the 12 selected countries are presented in Table S1. We observed that having the first ANC visit in the first trimester of pregnancy and at least two shots of tetanus toxoid were the components with lowest coverage for countries with high ANCq and low inequality, suggesting these are the last barriers to high ANCq scores. On the other extreme, among countries with below average coverage and high inequality, seeing a skilled provider and having the blood pressure measured were the interventions with higher coverage among the poorest. But the other interventions had low coverage, and especially the mean number of ANC visits was very low. The countries in the other two groups follow the same pattern, with higher coverage, generally, compared to the latter group. Among countries with the highest inequalities, having blood and urine samples collected seemed to be important drivers of inequality. The number of ANC visits also presented large differences between the richest and the poorest, several countries presenting differences around five visits. The most extreme example was India, with an SII of 5.6, and an average number of seven visits among the 20% richest women and 2.5 visits among the 20% poorest.

We also explored how the ANCq varied with woman’s empowerment measured through the three domains of the SWPER. Here we again found a systematic higher ANCq average score for women with higher levels of empowerment (Figure 4, and figures S1 and S2). The SWPER is available for a smaller number of countries, given it is estimable only for DHS surveys. Still, the widest gaps were found in countries from South Asia (notably Pakistan, India, and Nepal) and from East Asia and the Pacific (notably Myanmar).

**Figure 4.**
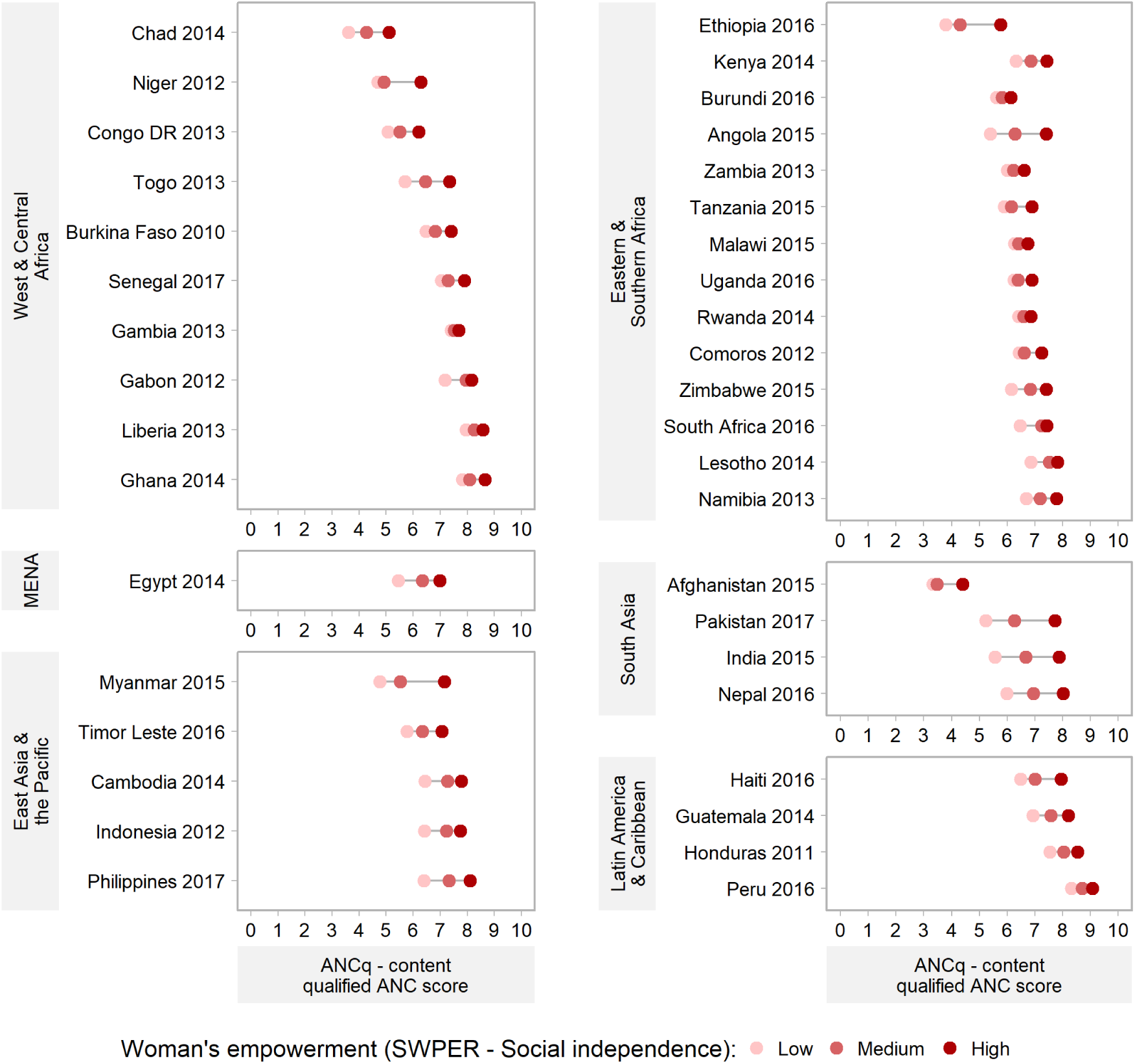
Equiplots of ANCq score by SWPER – Social Independence domain. Source: DHS, 2010-2017

Further stratified analyses showed that ANCq scores are systematically higher for women living in urban areas and with higher education level (Figures S3 and S4). A few countries did not present marked urban to rural differences, notably Thailand, Namibia, Eswatini, Maldives, El Salvador, despite having large rural populations. Clear patterns were not observed for woman’s age and sex of the child (Figures S5 and S6). Especially for sex of the child, the differences observed between girls and boys were very small, and no gender bias was evident an any of the countries studied, even in those where strong gender inequalities persist.

## Discussion

We explored socioeconomic inequalities in ANC in 63 LMICs using the ANCq indicator. Important inequalities in ANCq across socioeconomic groups were observed between and within countries and world regions. Women in urban areas, with secondary or more education, belonging to wealthier households and higher empowerment had higher ANCq scores in nearly all countries.

Studies measuring inequalities in maternal health care across a large number of countries have also shown that use of maternal health care varied greatly both within and between countries, and factors such as wealth, location, woman’s education, religion, and decision-making power are associated with the presence of inequalities.^1,2,9,21^ A study that analyzed 12 maternal, newborn and child health interventions from 54 countries found that four or more ANC visits was the second most inequitable indicator (after skilled attendant at birth), with an overall coverage of 49.5% (95%CI:35.6–66.7), and a difference of 34.6 percent points between women in the poorest quintile and those in the wealthiest.^2^

Several studies exploring the determinants on ANC consistently found that women living in urban areas, having higher levels of education, from the wealthiest households, and having higher empowerment levels are more likely to seek the recommended number of ANC visits, ensure early initiation or have good quality in ANC.^4,5,22–24^

A study conducted in São Tomé and Príncipe explored factors associated with adequate ANC found that it was adequate in 26% of the sample and was associated with maternal education and wealth. Women with higher education and belonging to the wealthiest households had four (OR:4.01; 95%CI:1.59–10.09) and two times (OR:1.99; 95%CI:1.19– 3.34) the odds of receiving adequate ANC compared to those with no education and women belonging to the poorest households, respectively.^25^

Similar findings were reported by Fagbamigbe and Idemudia^3^ in their study aimed to assess the quality of ANC services in Nigeria. Authors reported that less than 5% of ANC users received the desirable quality of ANC, and women with higher education (OR:2.69;95%CI:2.20–3.30), from wealthiest households (OR:3.54; 95%CI:2.65–4.72) had higher odds of receiving good quality in ANC; while women residing in rural areas (OR:0.83; 95%CI:0.74–0.94), and were not attended to by skilled ANC provider (OR:0.71; 95%CI:0.57–0.89) had lower odds.

Our findings are in line with the literature. Where our results advance the current knowledge is in the use of an indicator that includes information on quality and at the same time is applicable to all women in need of ANC. Our results show gaps that are not only related to having had contact with the services. When we find gaps wide as those presented by Angola and Pakistan, we see that the richest groups in those countries are on a par with the richest in the best performing countries, where women get a high number of visits and nearly all desired interventions. Furthermore, the poorest groups present scores that are among the worst, between 3 and 4 ANCq points. Women with 4 points in the ANCq score mostly had less than four ANC visits, tetanus immunization, a skilled provider, blood pressure measured, and nothing else.^8^

In terms of absolute wealth inequalities, measured by the SII, we observed that countries with higher mean ANCq presented lower inequalities, generally. Most of them are upper-middle income countries. Also, we noted that countries with average ANCq scores had a wide range of inequality, with some of them achieving very low inequality, as Malawi or Rwanda. Nevertheless, some countries showed high inequalities despite having average ANCq scores, such as India (ANCq=6.8; SII=4.6) or Pakistan (ANCq=6.5; SII=5.5). Countries with these characteristics are mostly low and lower-middle income countries. Our results also allow us to identify countries with very low ANCq, or very high inequality, or both. That can be a wake-up call for multilateral agencies and countries to focus their attention on this key aspect of maternal care. At the same time, we highlight some positive examples that could be studied and followed, like Thailand, Maldives and Dominican Republic.

Our results also showed that while there was large variability across countries in terms of mean ANCq, countries from Latin America and the Caribbean presented higher ANCq scores and less variability between them. In the same vein, an analysis of socioeconomic differences in the quality of ANC services in 59 LMICs from six world regions reported that Latin America and Caribbean women received more ANC services compared to women in the other regions.^21^ Additionally, a study conducted to analyze global inequality in maternal health care service utilization, mainly ANC and skilled birth assistance, showed that among the LMICs included, Latin America and Caribbean was the region with the highest prevalence of access to both care services, while Africa and Asia had the lowest prevalence and more disparities between countries,^9^ similar to our findings.

Monitoring health inequalities has become a priority in the Sustainable Development Goals (SDG) era helping countries to track progress towards the proposed goals and ensure that nobody is left behind.^26^ Despite all efforts in ANC programs, inequalities in coverage and quality of ANC services persist. Our findings suggest that interventions, that consider the social determinants of health and reduce socioeconomic inequalities in ANC are required in most LMICs. Also, those gaps that we documented must be bridged to achieve maternal and child mortality goals proposed in the 2030 SDG agenda.

Inequality is multidimensional, and disaggregating data permits tracking the health issues among disadvantaged subgroups considering contextual factors and priorities on a practical level.^27^ The information used in this paper is based on self-report, and it could be considered a limitation that should be noted, however all survey-based indicators used for SDG monitoring have the same problem.^27^ LMICs often lack good health information systems for monitoring health inequalities, and nationally representative surveys are, in most cases, the best available data source.^28^

Suitable approaches to monitoring ANC inequalities between and within countries are essential to provide evidence for practices, programs and policies aimed at reducing inequities,^28^ and to trace the impact of interventions. The ANCq is a new alternative, with several advantages, one of them being its ease of computation. It can be a valuable tool in this endeavor.

## Data Availability

The original datasets from Demographic and Health Survey (DHS) and Multiple Indicator Cluster Survey (MICS) are freely available.

http://dhsprogram.com/

http://mics.unicef.org/

## Acknowledgements

We thank the Bill & Melinda Gates Foundation, the Wellcome Trust, Associação Brasileira de Saúde Coletiva and Coordenação de Aperfeiçoamento de Pessoal de Nível Superior (CAPES) for funding this study. We are thankful to Thiago Melo for your help in the graphic design.

## Authors’ contributions

LA and AJDB conceptualized the paper and conducted the analyses, with support from CVG and GES. LA interpreted the results and wrote the manuscript with technical support from AJDB. AJDB, GES and CGV contributed to critically review the analysis and writing. All authors read and approved the final manuscript.

## Data availability statement

The original datasets from DHS (http://dhsprogram.com/) and MICS (http://mics.unicef.org/) are freely available.

## Open access

This is an open access article distributed in accordance with the Creative Commons Attribution Non Commercial (CC BY-NC 4.0) license, which permits others to distribute, remix, adapt, build upon this work non-commercially, and license their derivative works on different terms, provided the original work is properly cited, appropriate credit is given, any changes made indicated, and the use is non-commercial. See: http://creativecommons.org/licenses/by-nc/4.0/.

## References

1. Say L, Raine R. A systematic review of inequalities in the use of maternal health care in developing countries: examining the scale of the problem and the importance of context Public health reviews. Bull World Health Organ [Internet]. 2007 [cited 2020 Jan 21];85(10). Available from: http://www.who.

2. Barros AJ, Ronsmans C, Axelson H, Loaiza E, Bertoldi AD, Frana GV, et al. Equity in maternal, newborn, and child health interventions in Countdown to 2015: A retrospective review of survey data from 54 countries. Lancet [Internet]. 2012 Mar 31 [cited 2020 Sep 15];379(9822):1225–33. Available from: http://www.thelancet.com/article/S0140673612601135/fulltext

3. Fagbamigbe AF, Idemudia ES. Assessment of quality of antenatal care services in Nigeria: Evidence from a population-based survey [Internet]. Vol. 12, Reproductive Health. BioMed Central Ltd.; 2015 [cited 2020 Sep 15]. Available from: https://pubmed.ncbi.nlm.nih.gov/26382228/

4. Joshi C, Torvaldsen S, Hodgson R, Hayen A. Factors associated with the use and quality of antenatal care in Nepal: A population-based study using the demographic and health survey data. BMC Pregnancy Childbirth [Internet]. 2014 Mar 3 [cited 2020 Sep 15];14(1):94. Available from: http://bmcpregnancychildbirth.biomedcentral.com/articles/10.1186/1471-2393-14-94

5. Blackstone SR. Evaluating antenatal care in Liberia: evidence from the demographic and health survey. Women Heal [Internet]. 2019 Nov 26 [cited 2020 Sep 15];59(10):1141–54. Available from: https://pubmed.ncbi.nlm.nih.gov/30917774/

6. Kuhnt J, Vollmer S. Antenatal care services and its implications for vital and health outcomes of children: evidence from 193 surveys in 69 low-income and middle-income countries. BMJ Open. 2017 Nov;7(11):e017122.

7. Arsenault C, Jordan K, Lee D, Dinsa G, Manzi F, Marchant T, et al. Equity in antenatal care quality: an analysis of 91 national household surveys. Lancet Glob Heal. 2018 Nov;6(11):e1186–95.

8. Arroyave L, Saad GE, Victora CG, Barros AJD. A new content-qualified antenatal care coverage indicator: development and validation of a score using national health surveys. medRxiv. 2020 Feb 29;2020.02.28.20028720.

9. Yaya S, Ghose B. Global Inequality in Maternal Health Care Service Utilization: Implications for Sustainable Development Goals. [cited 2020 Jan 21]; Available from: http://online.liebertpub.com/doi/10.1089/heq.2018.0082

10. Corsi DJ, Neuman M, Finlay JE, Subramanian S V. Demographic and health surveys: a profile. Int J Epidemiol [Internet]. 2012;41(6):1602–13. Available from: http://files/6394/Corsi et al.- 2012 - Demographic and health surveys a profile.pdf

11. The DHS Program - What We Do [Internet]. [cited 2020 Mar 2]. Available from: https://dhsprogram.com/What-We-Do/index.cfm

12. Home - UNICEF MICS [Internet]. [cited 2020 Mar 2]. Available from: http://mics.unicef.org/

13. Indicators & Stratifiers-Int’l Center for Equity in Health [Internet]. [cited 2020 Jan 20]. Available from: http://equidade.org/indicators

14. Rutstein SO, Johnson K. The DHS wealth index [Internet]. DHS Comparative Reports No. 6.. Calverton, Maryland, USA: ORC Macro; 2004. Available from: http://dhsprogram.com/pubs/pdf/CR6/CR6.pdf

15. Filmer D, Pritchett L. Estimating Wealth Effects Without Expenditure Data--Or Tears: An Application to Educational Enrollments in States of India. Demography. 2001;38(1):115–32.

16. Rutstein SO. The DHS Wealth Index: Approaches for Rural and Urban Areas [Internet]. 2008 [cited 2020 Jan 20]. Available from: www.measuredhs.com

17. Ewerling F, Lynch JW, Victora CG, Van Eerdewijk A, Tyszler M, Barros AJD. The SWPER index for women’s empowerment in Africa: development and validation of an index based on survey data. 2017 [cited 2020 Jan 20]; Available from: www.thelancet.com/lancetgh

18. Ewerling F, Raj A, Victora CG, Hellwig F, Coll CVN, Barros A. A Survey-Based Women’s Empowerment Index for Low- and Middle-Income Countries: The SWPER Goes Global [Internet]. Rochester, NY: Social Science Research Network; 2019 Feb. Available from: https://papers.ssrn.com/abstract=3466986

19. World Bank. Population estimates and projections | DataBank [Internet]. [cited 2020 Sep 18]. Available from: https://databank.worldbank.org/reports.aspx?source=health-nutrition-and-population-statistics:-population-estimates-and-projections#

20. Barros AJD, Victora CG. Measuring Coverage in MNCH: Determining and Interpreting Inequalities in Coverage of Maternal, Newborn, and Child Health Interventions. [cited 2020 Jan 22]; Available from: www.plosmedicine.org

21. Amo-Adjei J, Aduo-Adjei K, Opoku-Nyamah C, Izugbara C. Analysis of socioeconomic differences in the quality of antenatal services in low and middle-income countries (LMICs). Grce M, editor. PLoS One [Internet]. 2018 Feb 23 [cited 2020 Sep 15];13(2):e0192513. Available from: https://dx.plos.org/10.1371/journal.pone.0192513

22. Luginaah IN, Kangmennaang J, Fallah M, Dahn B, Kateh F, Nyenswah T. Timing and utilization of antenatal care services in Liberia: Understanding the pre-Ebola epidemic context. Soc Sci Med [Internet]. 2016 Jul 1 [cited 2020 Sep 15];160:75–86. Available from: https://pubmed.ncbi.nlm.nih.gov/27214711/

23. Okonofua F, Ntoimo L, Ogungbangbe J, Anjorin S, Imongan W, Yaya S. Predictors of women’s utilization of primary health care for skilled pregnancy care in rural Nigeria. BMC Pregnancy Childbirth [Internet]. 2018 Apr 18 [cited 2020 Sep 15];18(1):106. Available from: https://bmcpregnancychildbirth.biomedcentral.com/articles/10.1186/s12884-018-1730-4

24. Saad-Haddad G, DeJong J, Terreri N, Restrepo-Méndez MC, Perin J, Vaz L, et al. Patterns and determinants of antenatal care utilization: Analysis of national survey data in seven countdown countries. J Glob Health [Internet]. 2016 [cited 2020 Sep 15];6(1). Available from: https://pubmed.ncbi.nlm.nih.gov/27231540/

25. dos Reis PADGD, Pereira CCDA, Leite I da C, Theme-Filha MM. Fatores associados à adequação do cuidado pré-natal e à assistência ao parto em São Tomé e Príncipe, 2008-2009. Cad Saude Publica [Internet]. 2015 Jan 1 [cited 2020 Sep 15];31(9):1929–40. Available from: http://dx.doi.org/10.1590/0102-311X00115914

26. Hosseinpoor AR, Bergen N, Schlotheuber A, Grove J. Measuring health inequalities in the context of sustainable development goals. Bull World Heal Organ [Internet]. 2018 [cited 2019 Aug 20];96(9):654–9. Available from: http://www.who.int/entity/bulletin/volumes/96/9/18-210401.pdf

27. Hosseinpoor AR, Bergen N, Koller T, Prasad A, Schlotheuber A, Valentine N, et al. Equity-Oriented Monitoring in the Context of Universal Health Coverage. PLoS Med. 2014 Sep 1;11(9).

28. Hosseinpoor AR, Bergen N, Schlotheuber A, Boerma T. National health inequality monitoring: current challenges and opportunities. Glob Health Action [Internet]. 2018 Dec 3 [cited 2020 Sep 12];11(sup1). Available from: /pmc/articles/PMC5827767/?report=abstract

