## Supplementary figures and images for "Inequalities in antenatal care coverage and quality: an analysis from 63 low and middle-income countries using the ANCq content-qualified coverage indicator"

### Figure S1

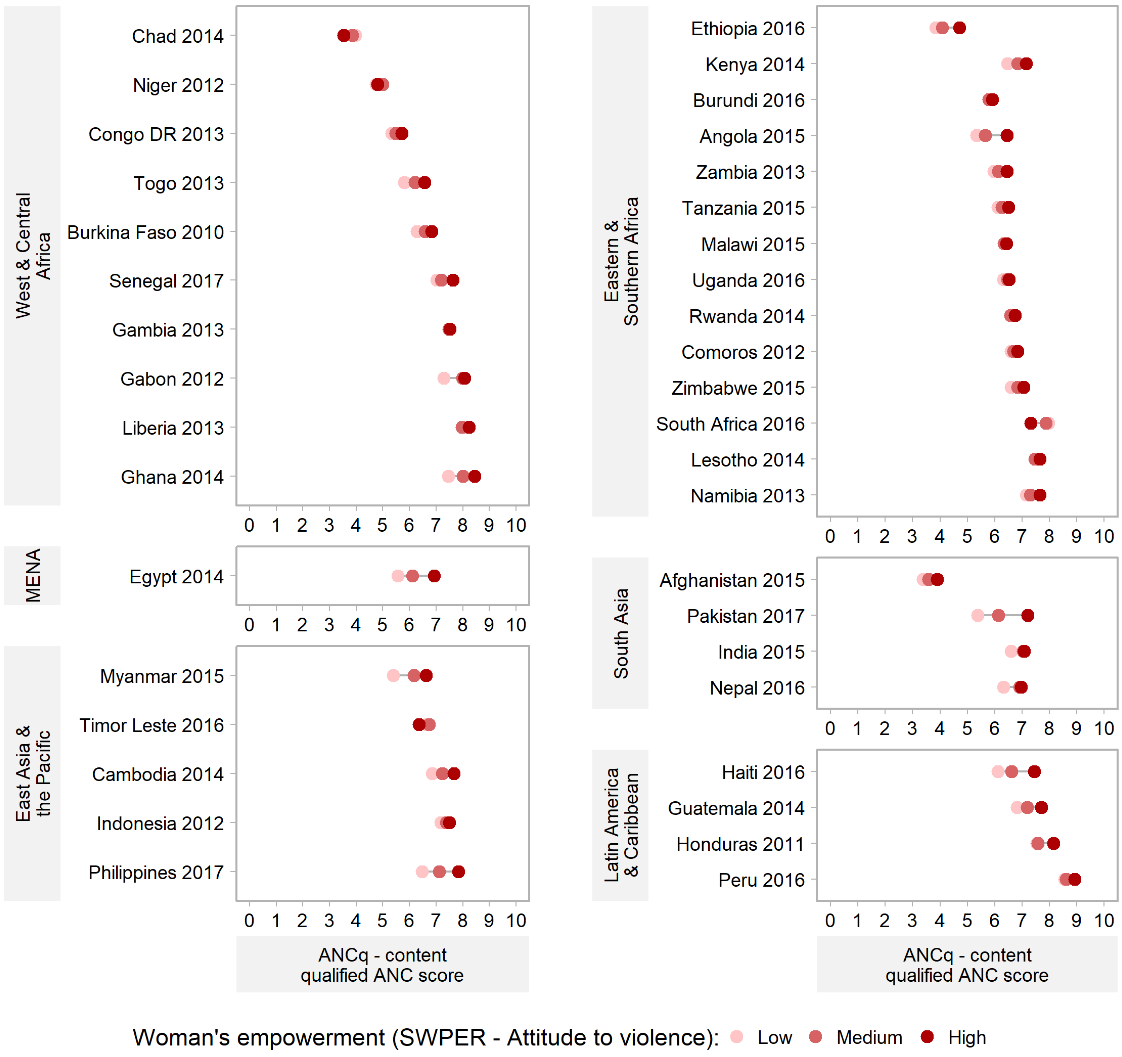


**Figure S1.** Equiplots of ANCq score by SWPER – Attitude to violence domain. Source: DHS, 2010-2017

### Figure S2

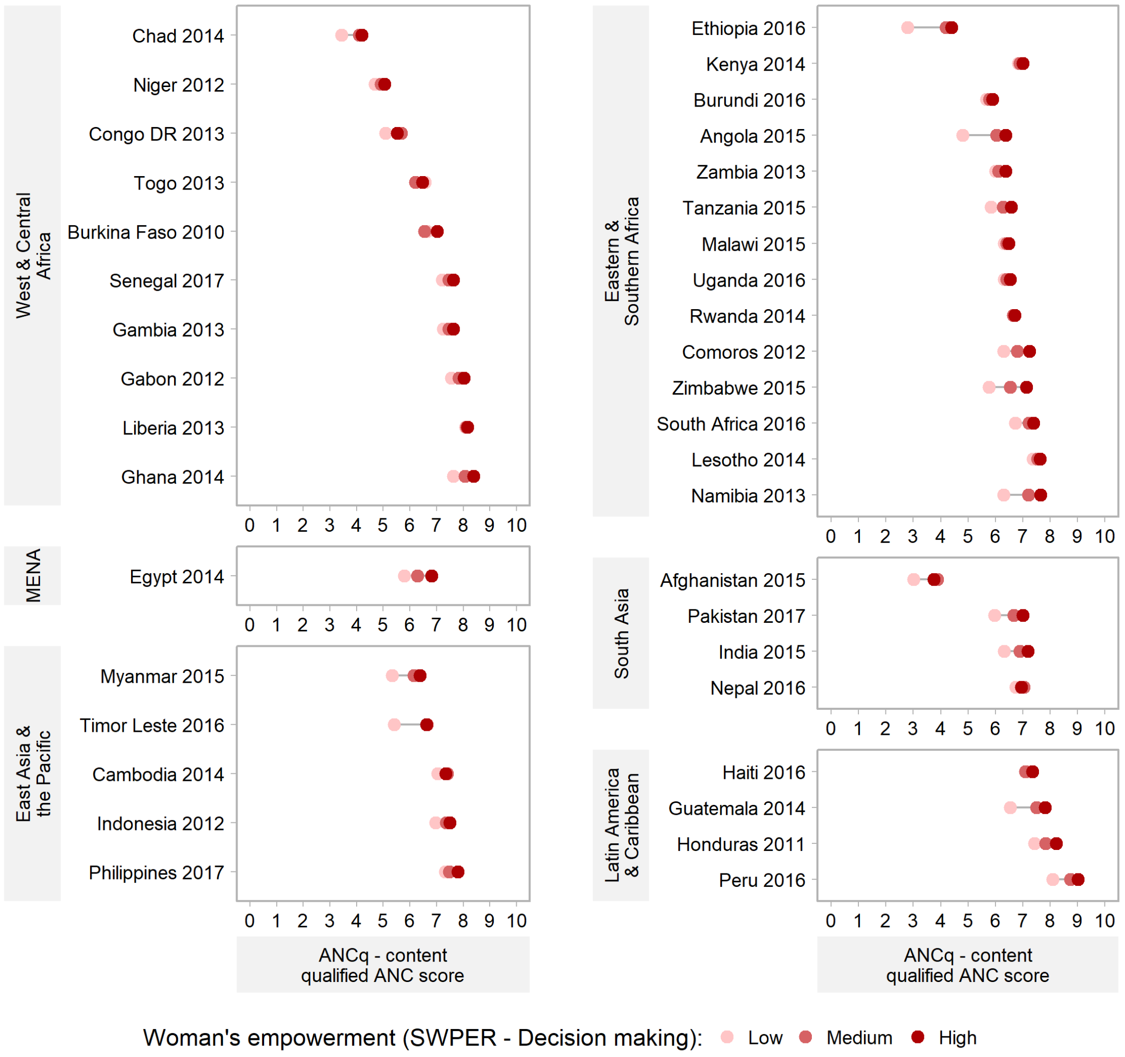


**Figure S2.** Equiplots of ANCq score by SWPER – Decision-making domain. Source: DHS, 2010-2017

### Figure S3

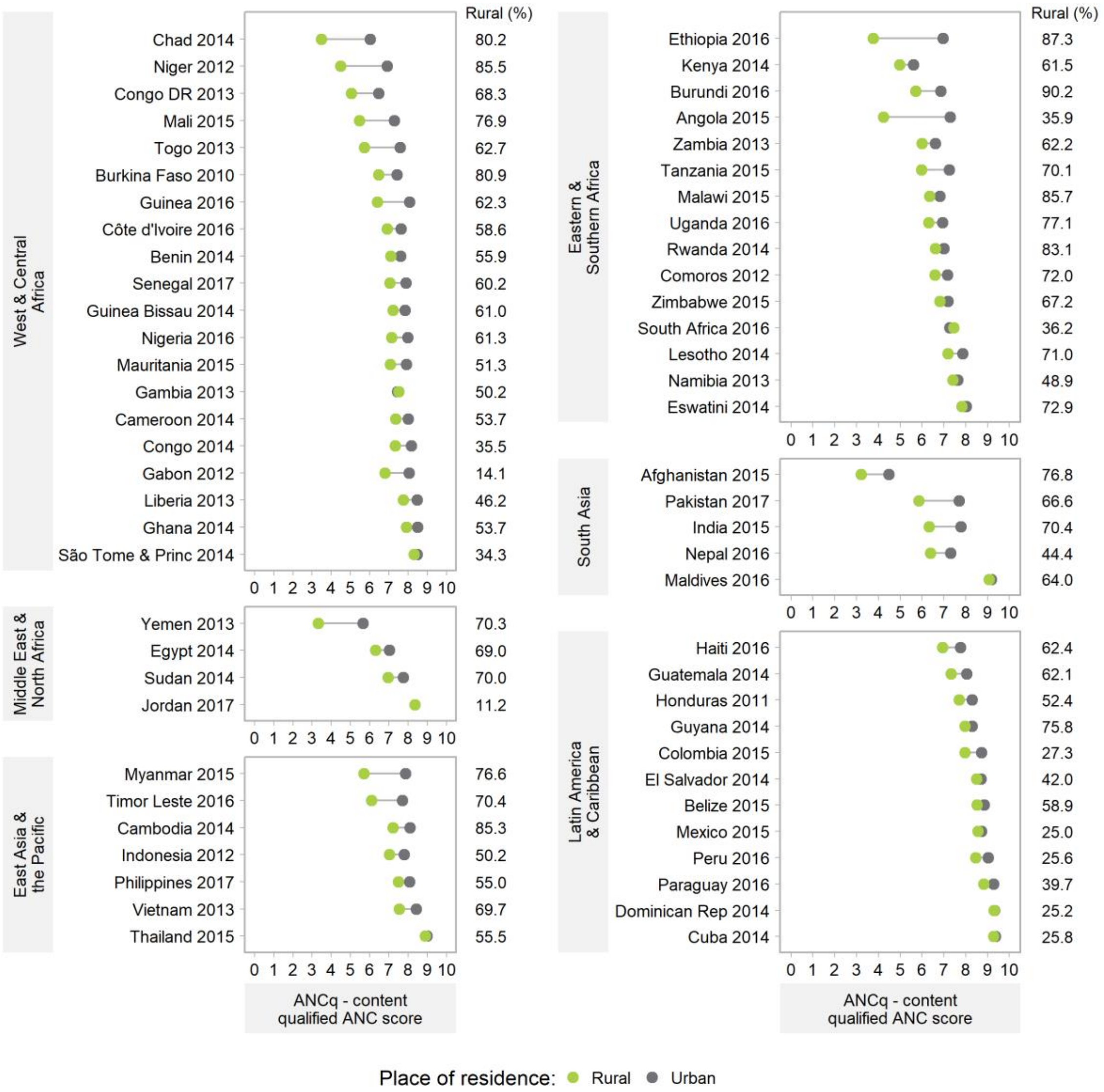


Figure S3. Equiplots of ANCq score by place of residence. Source: DHS and MICS, 2010-2017.

### Figure S4

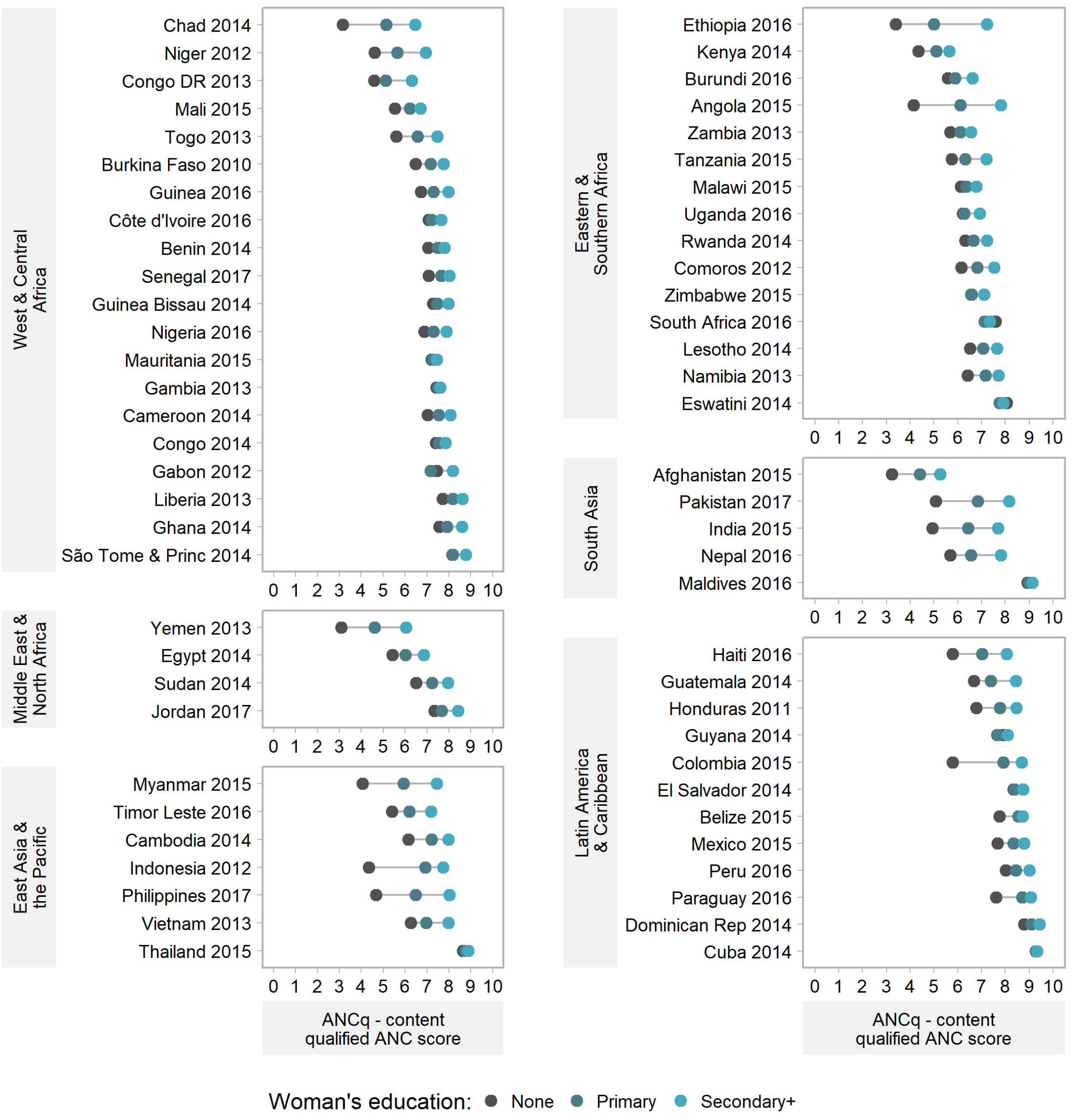


**Figure S4.** Equiplots of ANCq score by woman's education level. Source: DHS and MICS, 2010-2017.

### Figure S5

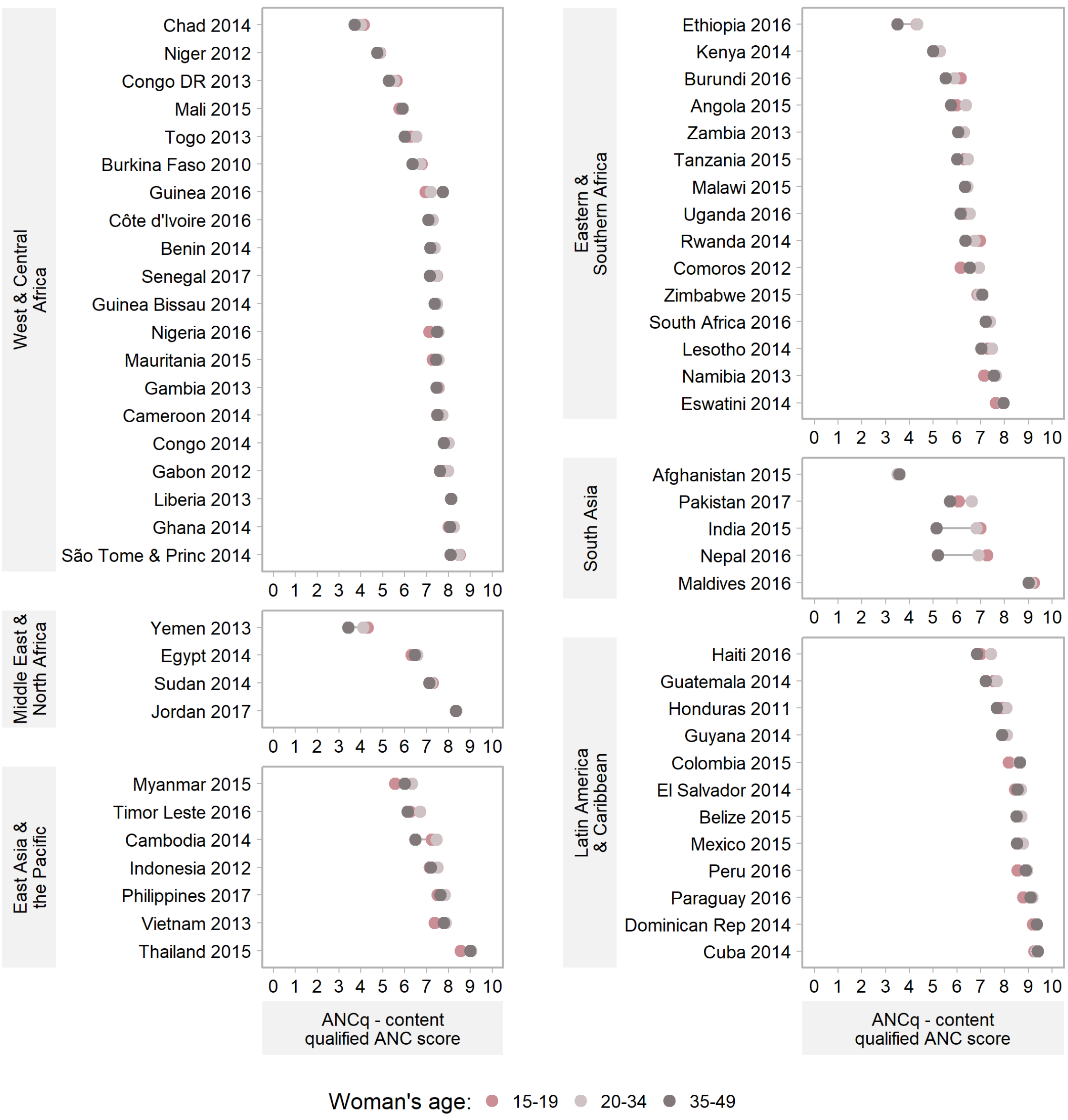


**Figure S5.** Equiplots of ANCq score by woman's age at childbirth. Source: DHS and MICS, 2010-2017.

### Figure S6

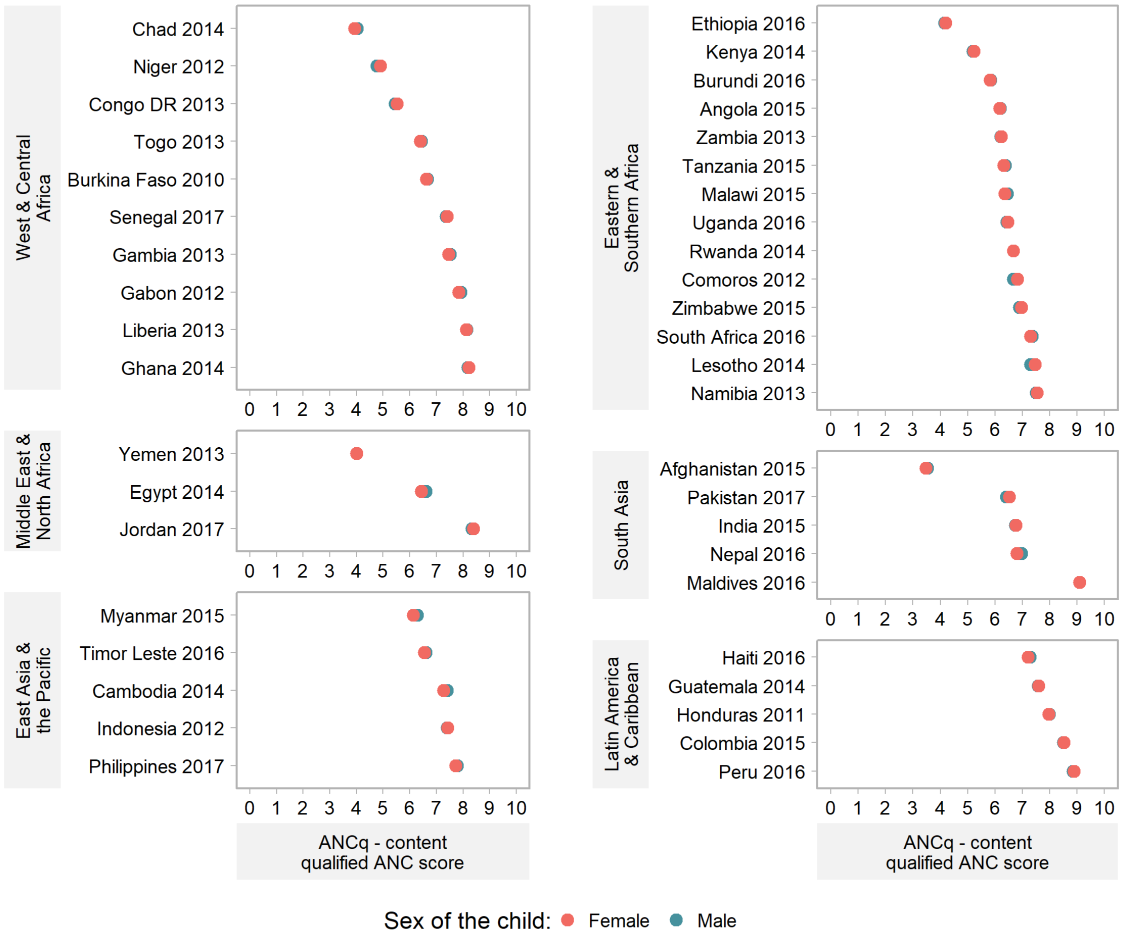


**Figure S6.** Equiplots of ANCq score by sex of the child. Source: DHS, 2010-2017.
