## Supplementary material for "Inequalities in antenatal care coverage and quality: an analysis from 63 low and middle-income countries using the ANCq content-qualified coverage indicator": Table S1

Table S1. Coverage and Slope Index of Inequality (absolute inequalities) of ANCq components for 12 LMICs. Source: DHS and MICS, 2010-2017.

| Country | Year | Wealth quintile | ANCq | ANCq components | | | | | | |
|---|---|---|---|---|---|---|---|---|---|---|
| | | | | ANC visits | Skilled | First trimester | Blood sample | Urine sample | Blood pressure | Tetanus |
| | | | Mean (SII) | Mean (SII) | % (SII) | % (SII) | % (SII) | % (SII) | % (SII) | % (SII) |
| Low and middle ANCq score and high inequality | | | | | | | | | | |
| Angola | 2015 | Q1 (poorest) | **3.8** | 2.4 | 55.0 | 24.2 | 36.9 | 32.0 | 42.2 | 35.3 |
| | | Q5 (wealthiest) | **8.3** | 5.9 | 98.0 | 66.6 | 96.2 | 95.8 | 95.3 | 77.5 |
| | | All | **6.2 (6.0)** | 4.1 (4.5) | 81.4 (54.6) | 40.1 (53.1) | 69.8 (81.5) | 67.3 (87.8) | 70.6 (71.1) | 56.1 (54.1) |
| Ethiopia | 2016 | Q1 (poorest) | **2.9** | 1.6 | 47.0 | 14.0 | 28.8 | 25.5 | 32.9 | 30.7 |
| | | Q5 (wealthiest) | **6.4** | 3.9 | 85.0 | 37.9 | 77.8 | 73.5 | 76.8 | 55.5 |
| | | All | **4.2 (3.7)** | 2.4 (2.5) | 62.3 (41.0) | 20.3 (23.5) | 45.6 (50.1) | 41.5 (50.4) | 47.3 (46.6) | 41.0 (28.6) |
| Afghanistan | 2015 | Q1 (poorest) | **2.9** | 1.4 | 50.0 | 22.2 | 7.9 | 13.4 | 42.8 | 33.1 |
| | | Q5 (wealthiest) | **4.8** | 2.9 | 76.0 | 44.7 | 32.5 | 37.4 | 62.5 | 33.8 |
| | | All | **3.5 (2.4)** | 1.9 (1.7) | 57.9 (32.2) | 30.1 (27.6) | 17.9 (29.8) | 24.0 (31.0) | 48.1 (25.4) | 33.3 (1.9) |
| High ANCq score and high inequality | | | | | | | | | | |
| India | 2015 | Q1 (poorest) | **4.7** | 2.5 | 57.0 | 37.6 | 45.5 | 47.3 | 49.2 | 78.2 |
| | | Q5 (wealthiest) | **8.4** | 7.0 | 94.1 | 77.3 | 92.2 | 91.8 | 92.9 | 87.6 |
| | | All | **6.8 (4.6)** | 4.8 (5.6) | 79.2 (46.1) | 58.5 (49.1) | 72.8 (58.0) | 73.3 (55.2) | 74.5 (54.3) | 83.0 (10.6) |
| Haiti | 2016 | Q1 (poorest) | **6.0** | 3.5 | 81.6 | 42.5 | 62.3 | 60.8 | 76.7 | 57.9 |
| | | Q5 (wealthiest) | **8.5** | 6.7 | 97.4 | 79.1 | 95.9 | 95.5 | 97.0 | 69.4 |
| | | All | **7.3 (3.0)** | 4.8 (3.6) | 91.0 (18.6) | 58.5 (44.6) | 80.4 (41.5) | 78.9 (41.5) | 88.2 (24.9) | 64.9 (13.5) |
| Vietnam | 2013 | Q1 (poorest) | **6.5** | 4.0 | 99.0 | 58.4 | 39.4 | 54.2 | 73.9 | 59.7 |
| | | Q5 (wealthiest) | **8.9** | 8.3 | 100.0 | 98.4 | 85.7 | 93.0 | 96.2 | 60.3 |
| | | All | **7.8 (2.8)** | 6.2 (5.4) | 99.8 (0.9) | 84.4 (45.3) | 64.3 (52.5) | 75.0 (38.6) | 85.7 (26.1) | 61.4 (4.1) |
| Low and middle ANCq score and low inequality | | | | | | | | | | |
| Kenya | 2014 | Q1 (poorest) | **4.6** | 3.3 | 89.7 | 13.0 | 38.9 | 32.7 | 37.9 | 20.1 |
| | | Q5 (wealthiest) | **5.8** | 4.9 | 99.0 | 31.3 | 45.6 | 45.4 | 45.7 | 29.3 |
| | | All | **5.2 (1.5)** | 4.0 (2.0) | 95.9 (10.6) | 19.2 (21.2) | 44.0 (8.5) | 40.7 (16.1) | 43.1 (10.7) | 24.3 (11.9) |
| Burundi | 2016 | Q1 (poorest) | **5.6** | 3.4 | 98.3 | 45.9 | 83.4 | 18.2 | 37.4 | 28.9 |
| | | Q5 (wealthiest) | **6.5** | 3.7 | 99.4 | 56.4 | 89.1 | 49.6 | 69.5 | 35.7 |
| | | All | **5.8 (0.9)** | 3.5 (0.2) | 99.2 (1.4) | 47.3 (6.9) | 85.6 (4.8) | 26.8 (31.3) | 47.4 (34.4) | 28.5 (5.6) |
| Malawi | 2015 | Q1 (poorest) | **6.2** | 3.6 | 93.1 | 21.5 | 89.0 | 28.9 | 78.0 | 72.6 |

| Country | Year | Wealth quintile | ANCq | ANCq components | | | | | | |
|---|---|---|---|---|---|---|---|---|---|---|
| | | | | ANC visits | Skilled | First trimester | Blood sample | Urine sample | Blood pressure | Tetanus |
| | | | Mean (SII) | Mean (SII) | % (SII) | % (SII) | % (SII) | % (SII) | % (SII) | % (SII) |
| | | Q5 (wealthiest) | **6.8** | 4.0 | 96.7 | 26.6 | 93.3 | 41.4 | 88.7 | 78.9 |
| | | All | **6.4 (0.6)** | 3.7 (0.4) | 94.8 (4.1) | 23.9 (5.9) | 90.8 (5.1) | 31.7 (12.6) | 81.7 (12.6) | 73.0 (3.5) |
| High ANCq score and low inequality | | | | | | | | | | |
| Dominican Rep | 2014 | Q1 (poorest) | **9.0** | 8.4 | 99.6 | 73.3 | 99.5 | 98.6 | 99.6 | 78.6 |
| | | Q5 (wealthiest) | **9.6** | 11.2 | 99.7 | 91.5 | 99.6 | 99.5 | 100.0 | 79.8 |
| | | All | **9.3 (0.7)** | 9.7 (3.3) | 99.6 (0.3) | 83.0 (22.2) | 99.7 (0.1) | 99.2 (0.9) | 99.8 (0.6) | 79.4 (2.3) |
| Jordan | 2017 | Q1 (poorest) | **8.1** | 7.8 | 96.4 | 84.2 | 90.8 | 90.0 | 92.2 | 10.5 |
| | | Q5 (wealthiest) | **8.5** | 9.8 | 96.5 | 83.6 | 95.2 | 94.9 | 95.9 | 15.6 |
| | | All | **8.4 (0.6)** | 8.8 (2.4) | 97.5 (1.3) | 84.9 (0.3) | 94.1 (6.7) | 93.2 (6.9) | 95.0 (5.4) | 10.0 (3.1) |
| Thailand | 2015 | Q1 (poorest) | **8.7** | 8.4 | 100.0 | 69.7 | 99.6 | 99.0 | 100.0 | 40.2 |
| | | Q5 (wealthiest) | **9.1** | 8.7 | 100.0 | 94.5 | 99.9 | 99.2 | 99.5 | 56.8 |
| | | All | **8.9 (0.4)** | 8.5 (0.0) | 100 (0.0) | 80.0 (29.8) | 99.7 (0.4) | 99.3 (-0.2) | 99.9 (-0.4) | 52.3 (17.8) |

SII: Slope Index of Inequality
