## Supplementary material for "Inequalities in antenatal care coverage and quality: an analysis from 63 low and middle-income countries using the ANCq content-qualified coverage indicator": Table S2

**Table S2.** Means and 95% confidence intervals of ANCq score by wealth quintiles. Source: DHS and MICS, 2010-2017.

| Country | Year | Source | Income Group | Wealth quintiles Q1 (Poorest) | Q2 | Q3 | Q4 | Q5 (Wealthiest) |
|---|---|---|---|---|---|---|---|---|
| | | | | ANCq Mean (95%CI) | ANCq Mean (95%CI) | ANCq Mean (95%CI) | ANCq Mean (95%CI) | ANCq Mean (95%CI) |
| West & Central Africa | | | | | | | | |
| Chad | 2014 | DHS | Low | 3.4 (3.1-3.7) | 3.5 (3.3-3.7) | 3.4 (3.2-3.6) | 3.5 (3.2-3.8) | 6.2 (6.0-6.3) |
| Niger | 2012 | DHS | Low | 3.8 (3.6-4.0) | 4.3 (4.1-4.5) | 4.6 (4.4-4.7) | 4.8 (4.6-5.1) | 6.5 (6.4-6.7) |
| Congo DR | 2013 | DHS | Low | 4.4 (4.2-4.7) | 5.0 (4.9-5.2) | 5.3 (5.1-5.5) | 5.8 (5.7-6.0) | 6.9 (6.7-7.0) |
| Mali | 2015 | MICS | Low | 5.0 (4.8-5.2) | 5.3 (5.1-5.4) | 5.3 (5.1-5.5) | 5.9 (5.7-6.1) | 7.4 (7.2-7.5) |
| Togo | 2013 | DHS | Low | 5.3 (5.1-5.6) | 5.4 (5.1-5.6) | 5.9 (5.7-6.1) | 7.3 (7.1-7.4) | 7.9 (7.8-8.0) |
| Burkina Faso | 2010 | DHS | Low | 5.9 (5.7-6.1) | 6.3 (6.2-6.4) | 6.5 (6.4-6.7) | 6.9 (6.8-7.0) | 7.4 (7.4-7.5) |
| Guinea | 2016 | MICS | Low | 6.0 (5.8-6.2) | 6.3 (6.1-6.4) | 6.8 (6.6-7.0) | 7.6 (7.4-7.8) | 8.3 (8.1-8.4) |
| Nigeria | 2016 | MICS | Lower-middle | 6.4 (6.2-6.6) | 6.8 (6.7-6.9) | 7.3 (7.2-7.4) | 7.7 (7.6-7.8) | 8.3 (8.2-8.3) |
| Senegal | 2017 | DHS | Low | 6.5 (6.4-6.7) | 7.0 (6.9-7.1) | 7.4 (7.3-7.5) | 7.8 (7.7-7.9) | 8.1 (8.1-8.2) |
| Benin | 2014 | MICS | Low | 6.6 (6.5-6.8) | 6.8 (6.7-7.0) | 7.2 (7.1-7.4) | 7.5 (7.3-7.6) | 8.2 (8.1-8.3) |
| Gabon | 2012 | DHS | Upper-middle | 6.7 (6.4-6.9) | 7.8 (7.6-8.0) | 8.0 (7.8-8.2) | 8.4 (8.1-8.6) | 8.3 (8.1-8.5) |
| Côte d'Ivoire | 2016 | MICS | Lower-middle | 6.7 (6.5-6.8) | 6.9 (6.7-7.0) | 7.1 (7.0-7.2) | 7.6 (7.5-7.8) | 8.0 (7.8-8.1) |
| Mauritania | 2015 | MICS | Lower-middle | 6.7 (6.5-6.9) | 7.0 (6.8-7.2) | 7.4 (7.2-7.5) | 7.7 (7.6-7.8) | 8.2 (8.1-8.3) |
| Cameroon | 2014 | MICS | Lower-middle | 7.0 (6.7-7.2) | 7.2 (7.1-7.4) | 7.6 (7.5-7.7) | 7.8 (7.8-7.9) | 8.3 (8.2-8.4) |
| Congo | 2014 | MICS | Lower-middle | 7.1 (7.0-7.2) | 7.7 (7.6-7.8) | 8.0 (7.9-8.1) | 8.2 (8.1-8.3) | 8.3 (8.2-8.5) |
| Guinea Bissau | 2014 | MICS | Low | 7.1 (7.0-7.2) | 7.1 (7.0-7.3) | 7.2 (7.1-7.4) | 7.7 (7.6-7.8) | 8.1 (7.9-8.3) |
| Ghana | 2014 | DHS | Lower-middle | 7.3 (7.0-7.7) | 7.8 (7.6-7.9) | 8.2 (8.1-8.4) | 8.5 (8.4-8.6) | 8.9 (8.8-9.0) |
| Liberia | 2013 | DHS | Low | 7.4 (7.2-7.6) | 7.7 (7.6-7.9) | 8.2 (8.0-8.3) | 8.6 (8.4-8.7) | 8.8 (8.6-9.0) |
| Gambia | 2013 | DHS | Low | 7.4 (7.3-7.5) | 7.5 (7.4-7.6) | 7.3 (7.2-7.5) | 7.4 (7.3-7.5) | 7.6 (7.5-7.8) |
| São Tome & Principe | 2014 | MICS | Lower-middle | 8.0 (7.7-8.2) | 8.3 (8.1-8.4) | 8.4 (8.2-8.6) | 8.5 (8.2-8.7) | 8.8 (8.5-9.0) |
| Eastern & Southern Africa | | | | | | | | |
| Ethiopia | 2016 | DHS | Low | 2.9 (2.6-3.2) | 3.5 (3.2-3.9) | 3.9 (3.6-4.2) | 4.5 (4.1-4.8) | 6.4 (6.1-6.7) |
| Angola | 2015 | DHS | Upper-middle | 3.7 (3.4-4.0) | 4.7 (4.5-4.9) | 6.7 (6.5-6.9) | 7.7 (7.6-7.9) | 8.3 (8.1-8.4) |
| Kenya | 2014 | DHS | Lower-middle | 4.5 (4.4-4.6) | 4.9 (4.8-5.0) | 5.1 (5.1-5.2) | 5.4 (5.3-5.5) | 5.7 (5.6-5.9) |
| Burundi | 2016 | DHS | Low | 5.5 (5.4-5.6) | 5.7 (5.6-5.7) | 5.7 (5.6-5.8) | 5.7 (5.6-5.8) | 6.5 (6.3-6.6) |
| Tanzania | 2015 | DHS | Low | 5.7 (5.5-5.8) | 5.8 (5.6-5.9) | 6.1 (6.0-6.2) | 6.6 (6.5-6.8) | 7.5 (7.3-7.6) |
| Zambia | 2013 | DHS | Lower-middle | 5.7 (5.6-5.8) | 5.9 (5.9-6.0) | 6.2 (6.1-6.2) | 6.4 (6.3-6.5) | 6.9 (6.8-7.0) |
| Comoros | 2012 | DHS | Low | 5.8 (5.4-6.2) | 6.3 (6.0-6.7) | 7.1 (6.8-7.4) | 7.1 (6.9-7.4) | 7.5 (7.2-7.8) |

| Country | Year | Source | Income Group | Wealth quintiles | | | | |
|---|---|---|---|---|---|---|---|---|
| | | | | Q1 (Poorest) | Q2 | Q3 | Q4 | Q5 (Wealthiest) |
| | | | | ANCq Mean (95%CI) | ANCq Mean (95%CI) | ANCq Mean (95%CI) | ANCq Mean (95%CI) | ANCq Mean (95%CI) |
| West & Central Africa | | | | | | | | |
| Uganda | 2016 | DHS | Low | 6.1 (6-6.2.0) | 6.1 (6.0-6.2) | 6.3 (6.2-6.4) | 6.5 (6.4-6.6) | 7.0 (6.9-7.1) |
| Malawi | 2015 | DHS | Low | 6.2 (6.1-6.3) | 6.3 (6.2-6.4) | 6.3 (6.2-6.3) | 6.4 (6.3-6.5) | 6.8 (6.7-6.9) |
| Zimbabwe | 2015 | DHS | Low | 6.5 (6.2-6.7) | 6.7 (6.5-7.0) | 6.9 (6.7-7.2) | 6.8 (6.7-7.0) | 7.6 (7.4-7.7) |
| Rwanda | 2014 | DHS | Low | 6.5 (6.4-6.6) | 6.5 (6.4-6.6) | 6.6 (6.5-6.7) | 6.6 (6.5-6.7) | 7.0 (6.9-7.1) |
| Lesotho | 2014 | DHS | Lower-middle | 6.8 (6.6-7.1) | 6.9 (6.7-7.2) | 7.2 (7.0-7.5) | 7.6 (7.4-7.8) | 8.1 (7.8-8.3) |
| South Africa | 2016 | DHS | Upper-middle | 7.1 (6.8-7.4) | 7.2 (6.8-7.5) | 7.3 (7.1-7.6) | 7.5 (7.3-7.7) | 7.4 (7.1-7.8) |
| Namibia | 2013 | DHS | Upper-middle | 7.2 (7.0-7.4) | 7.3 (7.1-7.5) | 7.5 (7.3-7.7) | 7.6 (7.5-7.8) | 7.8 (7.6-8.0) |
| Eswatini | 2014 | MICS | Lower-middle | 7.7 (7.6-7.8) | 7.7 (7.5-7.8) | 7.7 (7.5-7.9) | 7.9 (7.7-8.2) | 8.2 (7.9-8.4) |
| Middle East & North Africa | | | | | | | | |
| Yemen | 2013 | DHS | Lower-middle | 2.4 (2.2-2.7) | 2.8 (2.6-3.0) | 3.6 (3.4-3.8) | 4.8 (4.6-5.0) | 6.2 (6.0-6.4) |
| Egypt | 2014 | DHS | Lower-middle | 5.5 (5.3-5.7) | 6.0 (5.9-6.1) | 6.5 (6.4-6.6) | 7.0 (6.8-7.1) | 7.4 (7.3-7.6) |
| Sudan | 2014 | MICS | Lower-middle | 6.2 (6.0-6.4) | 6.7 (6.5-6.8) | 7.2 (7.0-7.3) | 7.5 (7.4-7.7) | 8.1 (8.0-8.2) |
| Jordan | 2017 | DHS | Upper-middle | 8.0 (7.9-8.1) | 8.3 (8.1-8.4) | 8.5 (8.4-8.6) | 8.5 (8.3-8.6) | 8.5 (8.2-8.8) |
| South Asia | | | | | | | | |
| Afghanistan | 2015 | DHS | Low | 2.8 (2.6-3.0) | 2.8 (2.6-3.1) | 3.2 (2.7-3.6) | 3.8 (3.6-4.1) | 4.7 (4.5-5.0) |
| Pakistan | 2017 | DHS | Lower-middle | 4.2 (3.9-4.5) | 5.3 (5.1-5.6) | 6.6 (6.4-6.9) | 7.7 (7.5-7.9) | 8.5 (8.4-8.6) |
| India | 2015 | DHS | Lower-middle | 4.6 (4.6-4.7) | 6.3 (6.2-6.3) | 7.2 (7.2-7.3) | 7.8 (7.7-7.8) | 8.3 (8.3-8.4) |
| Nepal | 2016 | DHS | Low | 5.7 (5.4-5.9) | 6.5 (6.2-6.7) | 6.8 (6.6-7.1) | 7.4 (7.2-7.6) | 8.2 (8.0-8.4) |
| Maldives | 2016 | DHS | Upper-middle | 9.0 (8.9-9.1) | 9.0 (8.8-9.1) | 9.1 (9.0-9.2) | 8.9 (8.7-9.1) | 9.3 (9.2-9.5) |
| East Asia & the Pacific | | | | | | | | |
| Myanmar | 2015 | DHS | Lower-middle | 4.7 (4.4-5.1) | 5.7 (5.4-6.0) | 6.3 (5.9-6.6) | 7.1 (6.8-7.3) | 8.2 (7.9-8.4) |
| Timor Leste | 2016 | DHS | Lower-middle | 5.4 (5.1-5.7) | 5.9 (5.6-6.1) | 6.2 (6.0-6.5) | 7.2 (7.0-7.4) | 7.9 (7.7-8.1) |
| Indonesia | 2012 | DHS | Lower-middle | 6.2 (6.0-6.3) | 7.2 (7.1-7.4) | 7.5 (7.4-7.6) | 7.9 (7.8-7.9) | 8.0 (8.0-8.1) |
| Vietnam | 2013 | MICS | Lower-middle | 6.5 (6.2-6.7) | 7.4 (7.1-7.6) | 7.6 (7.4-7.9) | 8.3 (8.1-8.5) | 8.8 (8.7-9.0) |
| Cambodia | 2014 | DHS | Low | 6.5 (6.2-6.8) | 7.0 (6.8-7.2) | 7.3 (7.1-7.5) | 7.7 (7.6-7.8) | 8.1 (8.0-8.2) |
| Philippines | 2017 | DHS | Lower-middle | 6.5 (6.3-6.8) | 7.4 (7.3-7.6) | 7.9 (7.8-8.1) | 8.5 (8.3-8.6) | 8.9 (8.7-9.0) |
| Thailand | 2015 | MICS | Upper-middle | 8.7 (8.5-8.9) | 8.8 (8.6-9.0) | 8.9 (8.8-9.1) | 9.0 (8.8-9.1) | 9.1 (8.9-9.2) |
| Latin America & Caribbean | | | | | | | | |
| Haiti | 2016 | DHS | Low | 5.9 (5.6-6.3) | 6.8 (6.5-7.0) | 7.4 (7.2-7.7) | 7.8 (7.6-7.9) | 8.5 (8.3-8.7) |
| Guatemala | 2014 | DHS | Lower-middle | 6.7 (6.5-6.9) | 7.2 (7.0-7.3) | 7.6 (7.4-7.7) | 8.2 (8.1-8.3) | 8.6 (8.5-8.7) |

| Country | Year | Source | Income Group | Wealth quintiles | | | | |
|---|---|---|---|---|---|---|---|---|
| | | | | Q1 (Poorest) | Q2 | Q3 | Q4 | Q5 (Wealthiest) |
| | | | | ANCq Mean (95%CI) | ANCq Mean (95%CI) | ANCq Mean (95%CI) | ANCq Mean (95%CI) | ANCq Mean (95%CI) |
| West & Central Africa | | | | | | | | |
| Honduras | 2011 | DHS | Lower-middle | 7.2 (7.1-7.3) | 7.6 (7.5-7.7) | 8.0 (7.9-8.2) | 8.3 (8.2-8.4) | 8.7 (8.6-8.8) |
| Guyana | 2014 | MICS | Lower-middle | 7.3 (7.1-7.5) | 8.1 (8.0-8.2) | 8.3 (8.1-8.5) | 8.3 (8.1-8.4) | 8.6 (8.4-8.8) |
| Colombia | 2015 | DHS | Upper-middle | 7.8 (7.7-8.0) | 8.4 (8.3-8.5) | 8.6 (8.5-8.7) | 8.8 (8.7-8.9) | 9.1 (8.9-9.3) |
| Belize | 2015 | MICS | Upper-middle | 8.2 (8.0-8.3) | 8.6 (8.3-8.8) | 8.7 (8.4-8.9) | 8.8 (8.7-9.0) | 9.0 (8.8-9.2) |
| Peru | 2016 | DHS | Upper-middle | 8.2 (8.1-8.4) | 8.8 (8.8-8.9) | 9.0 (8.9-9.0) | 9.1 (9.0-9.1) | 9.1 (9.1-9.2) |
| El Salvador | 2014 | MICS | Lower-middle | 8.3 (8.2-8.4) | 8.4 (8.3-8.5) | 8.7 (8.5-8.8) | 8.7 (8.6-8.8) | 8.9 (8.8-9.0) |
| Mexico | 2015 | MICS | Upper-middle | 8.4 (8.2-8.5) | 8.6 (8.4-8.7) | 8.7 (8.5-8.9) | 8.8 (8.7-9.0) | 9.0 (8.7-9.2) |
| Paraguay | 2016 | MICS | Upper-middle | 8.6 (8.4-8.7) | 8.9 (8.8-9.1) | 9.2 (9.1-9.3) | 9.3 (9.2-9.5) | 9.5 (9.4-9.6) |
| Dominican Rep | 2014 | MICS | Upper-middle | 9.0 (8.9-9.1) | 9.2 (9.1-9.3) | 9.4 (9.3-9.4) | 9.4 (9.4-9.5) | 9.5 (9.4-9.6) |
