## Supplementary material for "Inequalities in antenatal care coverage and quality: an analysis from 63 low and middle-income countries using the ANCq content-qualified coverage indicator": Table S5

**Table S5.** Means and 95% confidence intervals of ANCq score by SWPER - Decision-making domain. Source: DHS, 2010-2017.

| Country | Year | Income Group | Woman's Empowerment (SWPER – Decision Making) | | |
|---|---|---|---|---|---|
| | | | Low | Medium | High |
| | | | ANCq Mean (95%CI) | ANCq Mean (95%CI) | ANCq Mean (95%CI) |
| **West & Central Africa** | | | | | |
| Chad | 2014 | Low | 3.4 (3.3-3.5) | 4.1 (3.9-4.2) | 4.1 (3.9-4.3) |
| Niger | 2012 | Low | 4.6 (4.5-4.7) | 4.9 (4.8-5.0) | 5.0 (4.8-5.2) |
| Congo DR | 2013 | Low | 5.0 (4.9-5.2) | 5.6 (5.5-5.7) | 5.5 (5.4-5.6) |
| Togo | 2013 | Low | 6.5 (6.4-6.7) | 6.2 (6.0-6.3) | 6.4 (6.3-6.6) |
| Burkina Faso | 2010 | Low | 6.6 (6.5-6.7) | 6.5 (6.4-6.6) | 7.0 (6.9-7.1) |
| Senegal | 2017 | Low | 7.2 (7.1-7.2) | 7.4 (7.3-7.5) | 7.6 (7.5-7.7) |
| Gabon | 2012 | Upper-middle | 7.5 (7.2-7.9) | 7.8 (7.6-8.0) | 8.0 (7.8-8.2) |
| Gambia | 2013 | Low | 7.2 (7.0-7.4) | 7.4 (7.3-7.5) | 7.6 (7.5-7.6) |
| Liberia | 2013 | Low | 8.0 (7.8-8.3) | 8.0 (7.8-8.3) | 8.1 (8.0-8.2) |
| Ghana | 2014 | Lower-middle | 7.6 (7.3-7.9) | 8.0 (7.9-8.1) | 8.3 (8.3-8.4) |
| **Eastern & Southern Africa** | | | | | |
| Ethiopia | 2016 | Low | 2.7 (2.4-3.1) | 4.1 (3.9-4.4) | 4.3 (4.2-4.5) |
| Kenya | 2014 | Lower-middle | 6.8 (6.6-7.0) | 6.8 (6.8-6.9) | 7.0 (6.9-7.0) |
| Angola | 2015 | Upper-middle | 4.8 (4.3-5.2) | 6.0 (5.8-6.2) | 6.3 (6.2-6.5) |
| Burundi | 2016 | Low | 5.6 (5.5-5.7) | 5.7 (5.7-5.8) | 5.9 (5.8-5.9) |
| Zambia | 2013 | Lower-middle | 6.0 (5.8-6.1) | 6.1 (6.0-6.1) | 6.3 (6.3-6.4) |
| Tanzania | 2015 | Low | 5.8 (5.6-5.9) | 6.2 (6.1-6.3) | 6.5 (6.4-6.6) |
| Uganda | 2016 | Low | 6.3 (6.2-6.4) | 6.4 (6.3-6.4) | 6.5 (6.4-6.6) |
| Malawi | 2015 | Low | 6.3 (6.2-6.4) | 6.4 (6.3-6.4) | 6.4 (6.3-6.4) |
| Rwanda | 2014 | Low | 6.7 (6.5-6.8) | 6.6 (6.5-6.7) | 6.7 (6.6-6.7) |
| Comoros | 2012 | Low | 6.3 (6.0-6.5) | 6.7 (6.5-7.0) | 7.2 (6.9-7.5) |
| Zimbabwe | 2015 | Low | 5.7 (5.1-6.4) | 6.5 (6.3-6.7) | 7.1 (7.0-7.2) |
| Lesotho | 2014 | Lower-middle | 7.3 (6.9-7.8) | 7.5 (7.3-7.7) | 7.6 (7.5-7.7) |
| South Africa | 2016 | Upper-middle | 6.7 (5.9-7.4) | 7.2 (6.6-7.8) | 7.3 (7.2-7.5) |
| Namibia | 2013 | Upper-middle | 6.3 (5.4-7.1) | 7.2 (6.9-7.4) | 7.6 (7.5-7.7) |
| **Middle East & North Africa** | | | | | |
| Egypt | 2014 | Lower-middle | 5.7 (5.6-5.9) | 6.2 (6.1-6.3) | 6.8 (6.7-6.8) |
| **South Asia** | | | | | |
| Afghanistan | 2015 | Low | 3.0 (2.8-3.1) | 3.8 (3.7-4.0) | 3.7 (3.6-3.8) |
| Pakistan | 2017 | Lower-middle | 5.9 (5.8-6.1) | 6.6 (6.4-6.8) | 7.0 (6.8-7.1) |
| India | 2015 | Lower-middle | 6.3 (6.2-6.4) | 6.8 (6.7-6.9) | 7.1 (7.1-7.2) |
| Nepal | 2016 | Low | 6.7 (6.6-6.8) | 7.0 (6.8-7.2) | 6.9 (6.7-7.1) |
| **East Asia & the Pacific** | | | | | |
| Myanmar | 2015 | Lower-middle | 5.3 (4.8-5.8) | 6.1 (5.9-6.3) | 6.3 (6.2-6.5) |
| Timor Leste | 2016 | Lower-middle | 5.4 (4.6-6.1) | 6.6 (6.3-6.9) | 6.6 (6.5-6.7) |
| Indonesia | 2012 | Lower-middle | 6.9 (6.7-7.1) | 7.3 (7.2-7.4) | 7.4 (7.4-7.5) |
| Cambodia | 2014 | Low | 7.0 (6.6-7.4) | 7.4 (7.2-7.5) | 7.3 (7.2-7.4) |
| Philippines | 2017 | Lower-middle | 7.3 (6.9-7.6) | 7.4 (7.2-7.6) | 7.8 (7.7-7.8) |
| **Latin America & Caribbean** | | | | | |
| Haiti | 2016 | Low | 7.2 (6.9-7.5) | 7.1 (6.9-7.2) | 7.3 (7.2-7.4) |
| Guatemala | 2014 | Lower-middle | 6.5 (6.2-6.8) | 7.5 (7.3-7.6) | 7.8 (7.7-7.8) |
| Honduras | 2011 | Lower-middle | 7.4 (7.1-7.6) | 7.8 (7.7-7.9) | 8.2 (8.1-8.2) |
| Peru | 2016 | Upper-middle | 8.1 (7.8-8.3) | 8.7 (8.6-8.8) | 9.0 (8.9-9.0) |
